# Evaluating the effect of metabolic traits on oral and oropharyngeal cancer risk using Mendelian randomization

**DOI:** 10.1101/2022.08.10.22278617

**Authors:** Mark Gormley, Tom Dudding, Steven J Thomas, Jessica Tyrrell, Andrew R Ness, Miranda Pring, Danny Legge, George Davey Smith, Rebecca C Richmond, Emma E Vincent, Caroline Bull

## Abstract

A recent World Health Organization report states that at least 40% of all cancer cases may be preventable, with smoking, alcohol consumption and obesity identified as three of the most important modifiable lifestyle factors. Given the significant decline in smoking rates, particularly within developing countries, other potentially modifiable risk factors for head and neck cancer warrant investigation. Obesity and related metabolic disorders such as type 2 diabetes and hypertension have been associated with head and neck cancer risk in multiple observational studies. However, obesity has also been correlated with smoking, with bias, confounding or reverse causality possibly explaining these findings. To overcome the challenges of observational studies, we conducted two-sample Mendelian randomization (inverse variance weighted (IVW) method) using genetic variants which were robustly associated with obesity, T2D and hypertension in genome-wide association studies (GWAS). Outcome data was taken from the largest available GWAS of 6,034 oral and oropharyngeal cases, with 6,585 controls. We found limited evidence of a causal effect of genetically proxied body mass index (OR IVW = 0.89, 95%CI 0.72–1.09, p = 0.26 per 1 SD in BMI (4.81 kg/m2)) on oral and oropharyngeal cancer risk. Similarly, there was limited evidence for related traits including type 2 diabetes and hypertension. Smoking appears to act as a mediator in the relationship between obesity and head and neck cancer.

## Introduction

Head and neck squamous cell carcinoma (HNC), which includes cancers of the oral cavity and oropharynx is the 7^th^ most common cancer, accounting for more than 660,000 new cases and 325,000 deaths annually worldwide (Johnson et al., 2020; Sung et al., 2021). Established risks include tobacco use, alcohol consumption (Hashibe et al., 2009) and human papillomavirus (HPV) infection, mainly associated with oropharyngeal cancer and thought to be sexually transmitted (Gillison, Chaturvedi, Anderson, & Fakhry, 2015). A recent World Health Organization (WHO) report states that at least 40% of all cancer cases may be preventable, with smoking, alcohol consumption and obesity identified as three of the most important modifiable lifestyle factors (World Health Organization (WHO), 2022). Smoking behaviour is declining, particularly in developing countries and it has been projected that obesity could even supersede smoking as the primary driver of cancer in the coming decades (World Health Organization (WHO), 2022). Despite changes in smoking rates, the incidence of HNC continues to rise and a changing aetiology has been proposed (Conway, 2018; Thomas, Penfold, Waylen, & Ness, 2018). Therefore, less established risks such as obesity and its related metabolic traits warrant investigation in HNC. However, obesity has been correlated with other HNC risk factors such as smoking (Carreras-Torres et al., 2018), alcohol (Alice R. Carter et al., 2019) and educational attainment (A. R. Carter et al., 2019), meaning independent effects are difficult to establish.

Obesity is now considered to increase the risk of at least 13 different types of cancer including breast, colorectal, gastric and oesophageal (Centers for Disease Control and Prevention, 2021), but the effect on HNC risk remains unclear (World Health Organization (WHO), 2022). Public health strategies have been unsuccessful in addressing the current obesity epidemic at the population level, which could result in more cancer cases in the years to come (Davey, 2004). Obesity and related metabolic traits such as type 2 diabetes (T2D), hypertension and dyslipidaemia have all been associated with head and neck cancer in multiple observational studies. In the largest pooled analysis, obesity defined by higher body mass index (BMI) was associated with a protective effect for HNC in current smokers (hazard ratio (HR)10.76, 95% confidence intervals (95%CI) 0.71–0.82, *p*⍰<0.0001, per 5⍰kg/m^2^) and conversely, a higher risk in never smokers (HR 1.15, 95%CI 1.06–1.24 per 5⍰kg/m^2^, *p* < 0.001) (Gaudet et al., 2015). In the same study, a greater waist circumference (WC) (HR⍰ ⍰1.04, 95%CI 1.03–1.05 per 5⍰cm, *p*⍰< 0.001) and waist-to-hip ratio (WHR) (HR⍰1.07, 95%CI 1.05–1.09 per 0.1 unit, *p* < 0.001) were associated with increased HNC risk, which did not vary by smoking status (Gaudet et al., 2015). However, more recent cohort studies have failed to show a clear association between BMI and HNC (Cao et al., 2020; Diaz et al., 2021; Gribsholt et al., 2020; Jiang et al., 2021; Ward et al., 2017). A meta-analysis of observational studies investigating T2D with oral and oropharyngeal subsites, showed an increased risk ratio (RR) of 1.15, 95%CI 1.02–1.29, *P heterogeneity* = 0.277 (Gong, Wei, Yu, & Pan, 2015), a result which is consistent with more recent independent cohorts (Jiang et al., 2021; H.-B. Kim, Kim, Han, & Joo, 2021; S.-Y. Kim, Han, & Joo, 2019; Saarela et al., 2019). Hypertension (defined as a systolic blood pressure (SBP) > 130 mmHg or diastolic blood pressure (DBP) > 85 mmHg), has been correlated with head and neck cancer risk across multiple studies (Christakoudi et al., 2020; H.-B. Kim et al., 2021; S.-Y. Kim et al., 2019; Seo, Kim, Park, Han, & Joo, 2020; Stocks et al., 2012). Nonetheless, selection bias, confounding, or reverse causation may explain the findings from these studies.

Mendelian randomization (MR) is an analytical approach which attempts to overcome the challenges of conventional epidemiological studies. The method uses germline genetic single nucleotide polymorphisms (SNPs), which are randomly assorted during meiosis (and fixed at conception), to estimate the causal effects of exposures on disease outcomes (Davey Smith & Hemani, 2014; Sanderson et al., 2022; Smith & Ebrahim, 2003). Using MR, we recently found limited evidence for a role of circulating lipid traits in oral and oropharyngeal cancer risk (Gormley et al., 2021), however other metabolic traits remain untested in an MR framework. Here, we employ a two-sample MR approach, integrating summary-level genetic data from the largest available GWAS for metabolic traits, including obesity measures (BMI, WC, WHR), glycaemic traits (T2D, glycated haemoglobin (HbA_1c_), fasting glucose (FG), fasting insulin (FI)), and blood pressure (SBP, DBP) to evaluate their causal effect on oral and oropharyngeal cancer risk. Given the potential correlation of metabolic traits and established HNC risk factors, further evaluation of instrument-risk factor effects including smoking, alcohol, risk tolerance (as a proxy for sexual behaviour), and educational attainment was carried out using MR.

## Methods

Two-sample MR was performed using published summary-level data from the largest available GWAS for each metabolic trait. MR makes three key assumptions, as described in **Figure 1** (Davey Smith & Hemani, 2014; Smith & Ebrahim, 2003).

**Figure 1.**
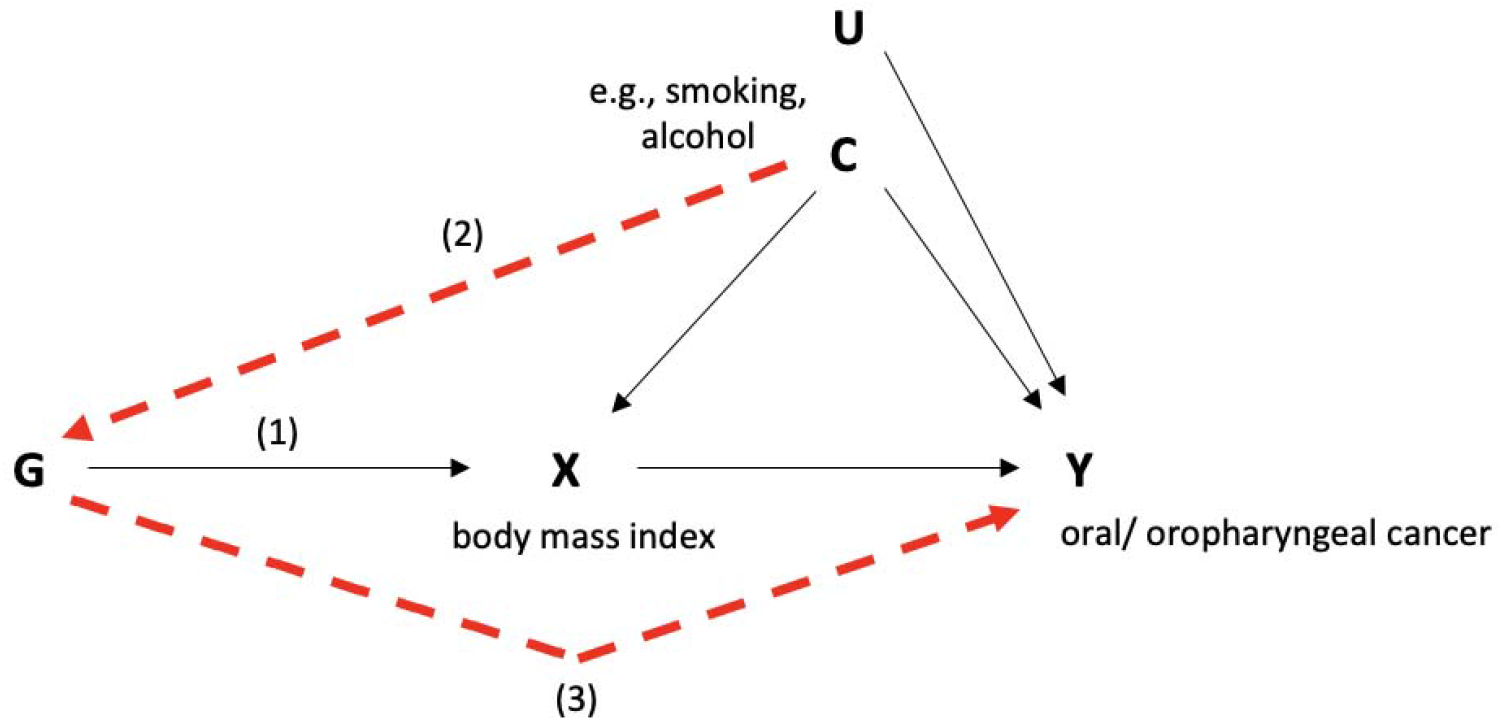
Directed acyclic graph (DAG) depicting Mendelian randomization applied to this study Genetic variants (G) act as proxies or instruments to investigate if an exposure (X), is associated with a trait e.g., body mass index or disease outcome (Y) e.g., oral/oropharyngeal cancer. Causal inference can be made between X and Y if the following conditions are upheld: (1)G is a valid instrument, reliably associated with X (‘relevance’); (2)no measured (C) or unmeasured (U) confounding of the association between G and the Y (‘exchangeability’); (3)there is no independent association of G with Y, except through X (‘exclusion restriction principle’).

### Exposure summary statistics for metabolic traits

To instrument metabolic traits, we selected genetic variants associated (*p* < 5 × 10^−8^) with traits of interest identified by previously conducted GWAS **(Supplementary file 1)**. Clumping was performed to ensure single nucleotide polymorphisms (SNPs) in each instrument were independent (linkage disequilibrium R^2^ < 0.001). Following clumping, genetic instruments were comprised of: 312 SNPs for BMI, from a GWAS meta-analysis of 806,834 individuals of European ancestry, including the Genetic Investigation of ANthropometric Traits (GIANT) consortium and UK Biobank (Pulit et al., 2019); and 209 SNPs for WHR extracted from the same GWAS in 697,734 individuals (Pulit et al., 2019). 45 SNPs for WC were taken from a GWAS meta-analysis describing 224,459 individuals of mainly European ancestry (Shungin et al., 2015); 275 SNPs for T2D from the DIAMANTE (DIAbetes Meta-ANalysis of Trans-Ethnic association studies) consortium of 228,499 cases and 1,178,783 controls (Vujkovic et al., 2020); 33 SNPs for FG and 18 SNPs for FI, obtained from a GWAS published by the MAGIC (Meta-Analyses of Glucose and Insulin-Related Traits) Consortium (N = 151,188 and 105,056 individuals, respectively) (Lagou et al., 2021); 58 SNPs for HbA_1c_, taken from a meta-analyses of 159,940 individuals from 82 cohorts of European, African, East Asian, and South Asian ancestry (Wheeler et al., 2017); Finally, 105 and 78 SNPs for SBP and DBP, respectively, were extracted from a GWAS meta-analysis of over 1 million participants in UK Biobank and the International Consortium of Blood Pressure Genome Wide Association Studies (ICBP) (Evangelou et al., 2018) **(Supplementary file 1)**.

### Outcome summary statistics for oral and oropharyngeal cancer

We estimated the effects of metabolic traits on risk of oral and oropharyngeal cancer by extracting exposure SNPs **(Supplementary file 1)** from the largest available GWAS performed on 6,034 cases and 6,585 controls from 12 studies which were part of the Genetic Associations and Mechanisms in Oncology (GAME-ON) Network (Lesseur et al., 2016). Full details of the included studies, as well as the genotyping and imputation performed, have been described previously (Dudding et al., 2018; Lesseur et al., 2016). In brief, the study population included participants from Europe (45.3%), North America (43.9%) and South America (10.8%). Cancer cases comprised the following the International Classification of Diseases (ICD) codes: oral cavity (C02.0-C02.9, C03.0-C03.9, C04.0-C04.9, C05.0-C06.9) oropharynx (C01.9, C02.4, C09.0-C10.9), hypopharynx (C13.0-C13.9), overlapping (C14 and combination of other sites) and 25 cases with unknown ICD code (other). A total of 954 individuals with cancers of hypopharynx, unknown code or overlapping cancers were excluded. Genomic DNA isolated from blood or buccal cells was genotyped at the Center for Inherited Disease Research (CIDR) using an Illumina OncoArray, custom designed for cancer studies by the OncoArray Consortium (Consortium, 2013). Principle components analysis was performed using approximately 10,000 common markers in low LD (r^2^ < 0.004), minor allele frequency (MAF) > 0.05 and 139 population outliers were removed.

Given the differential association of potential risk factors at each subsite (i.e. smoking, alcohol and HPV infection)(Thomas et al., 2018), we performed stratified MR analyses for oral and oropharyngeal cancer to evaluate potential heterogeneity in effects. For this, we used GWAS summary data on a subset of 2,990 oral and 2,641 oropharyngeal cases and the 6,585 common controls in the GAME-ON GWAS (Lesseur et al., 2016).

### Statistical analysis

Two-sample MR was conducted using the “TwoSampleMR” package in R (version 3.5.3), by integrating SNP associations for each metabolic trait (exposure, sample 1) with those for oral and oropharyngeal cancer in GAME-ON (outcome, sample 2). We only used genetic variants reaching GWAS significance (*p* <5×10^−8^). The nearest gene was identified using SNPsnap and a distance of +/- 500 kb (Pers, Timshel, & Hirschhorn, 2014). Firstly, metabolic trait-associated SNPs were extracted from oral and oropharyngeal cancer summary statistics. Exposure and outcome summary statistics were harmonised using the & *#x201C;harmonise_data”* function of the TwoSampleMR package so that variant effect estimates corresponded to the same allele. Palindromic SNPs were identified and corrected using allele frequencies where possible (alleles were aligned when minor allele frequencies were < 0.3, or were otherwise excluded). For each SNP in each exposure, individual MR effect-estimates were calculated using the Wald method (SNP-outcome beta/SNP-exposure beta) (Wald, 1940). Multiple SNPs were then combined into multi-allelic instruments using random-effects inverse-variance weighted (IVW) meta-analysis.

IVW estimates may be vulnerable to bias if genetic instruments are invalid and are only unbiased in the absence of horizontal pleiotropy or when horizontal pleiotropy is balanced (Hemani, Bowden, & Smith, 2018). We therefore performed additional sensitivity analyses to evaluate the potential for unbalanced horizontal pleiotropy using weighted median (Bowden, Smith, Haycock, & Burgess, 2016), weighted mode (Hartwig, Smith, & Bowden, 2017) and MR-Egger (Bowden, Davey Smith, & Burgess, 2015) methods which are described in detail elsewhere (Lawlor DA, 2019). In short, the weighted median stipulates that at least 50% of the weight in the analysis stems from valid instruments. Weighted mode returns an unbiased estimate of the causal effect if the cluster with the largest weighted number of SNPs for the weighted model are all valid instruments. Instruments are weighted by the inverse variance of the SNP-outcome association (Hartwig et al., 2017).

Finally, MR-Egger provides reliable effect estimates even if variants are invalid and the Instrument Strength Independent of Direct Effect (InSIDE) assumption is violated (Bowden et al., 2015). The InSIDE assumption states that the association between genetic instrument and exposure should not be correlated with an independent path from instrument to the outcome. In the presence of unbalanced pleiotropy when the InSIDE assumption is violated, then the MR-Egger result may be biased (Lawlor DA, 2019). Gene variants must be valid instruments and where there was evidence of violation of the negligible measurement error (NOME) assumption (Bowden, Del Greco, et al., 2016), this was assessed using the I^2^ statistic and MR-Egger was performed with simulation extrapolation (SIMEX) correction for bias adjustment (Bowden, Del Greco, et al., 2016). The variance of each trait explained by the genetic instrument (*R*^*2*^) was estimated and used to perform power calculations (Brion, Shakhbazov, & Visscher, 2013). F-statistics were also generated. An F-statistic lower than 10 was interpreted as indicative of a weak instrument bias (Lawlor, Harbord, Sterne, Timpson, & Davey Smith, 2008). To further assess the robustness of MR estimates, we examined evidence of heterogeneity across individual SNPs using the Cochran Q-statistic, which indicates the presence of invalid instruments (e.g., due to horizontal pleiotropy), if Q is much larger than its degrees of freedom (No. of instrumental variables minus 1) (Bowden et al., 2018). MR-PRESSO (Mendelian Randomization Pleiotropy RESidual Sum and Outlier) was used to detect and correct for potential outliers (where Q-statistic p < 0.05) (Verbanck, Chen, Neale, & Do, 2018).

### Instrument-risk factor effects

Where there was evidence for an effect of a metabolic trait on oral or oropharyngeal cancer risk in the primary MR analysis, we conducted further evaluation of the metabolic instruments onto established HNC risk factors using two-sample MR. The largest available GWAS were used for smoking initiation (a binary phenotype indicating whether an individual had ever smoked in their life versus never smokers) (n=⍰1,232,091) and alcoholic drinks per week (defined as the average number of drinks per week aggregated across all types of alcohol, n=⍰941,280) from the GWAS and Sequencing Consortium of Alcohol and Nicotine use (GSCAN) study (Liu et al., 2019). Summary statistics were also obtained from a GWAS of general risk tolerance (n=⍰939,908), derived from a meta-analysis of UK Biobank (n=⍰431,126) binary *question “Would you describe yourself as someone who takes risks⁵”* and the 23andMe (n=⍰508,782) question *“Overall, do you feel comfortable or uncomfortable taking risks⍰”*. The GWAS of risk tolerance was based on one’s tendency or willingness to take risks, making them more likely to engage in risk-taking behaviours more generally (Karlsson Linner et al., 2019). A strong genetic correlation between sexual behaviours and risk tolerance has been shown previously (Gormley et al., 2022). Finally, given the known association between HNC and lower socioeconomic position, we used MR to examine educational attainment (defined by years of schooling) (J. Lee et al., 2018). Outcome beta estimates reflect the standard deviation of the phenotype.

## Results

F-statistics of genetic instruments for metabolic traits ranged from 33.3 – 133.6, indicating sufficient instrument strength for MR analyses **(Supplementary file 2 – table S1)**. Genetic instruments were estimated to explain between 0.5% (FI) and 4% (BMI) of their respective metabolic trait (**Supplementary file 2 – table S1**). Based on the results of prior observational studies we would expect to detect OR of >1.10 for a clinically meaningful effect of metabolic traits on oral and oropharyngeal cancer. **Supplementary file 2 – figure S1A-C** displays power estimates for MR analyses. In analyses where BMI was the exposure, we had 70% power to detect an association with an OR of 1.2 or more at an α of 0.05 for combined oral and oropharyngeal cancer. Power was lower for other metabolic traits and reduced when stratifying analyses by subsite **(Supplementary file 2 – figure S1B-C)**.

### Estimated effect of adiposity on oral and oropharyngeal cancer risk

There was limited evidence of an effect of higher BMI or WHR on combined oral and oropharyngeal cancer (OR IVW = 0.89, 95%CI 0.72–1.09, *p* = 0.26, per 1 SD in BMI (4.81 kg/m^2^) and OR IVW = 0.98, 95%CI 0.74–1.29, *p* = 0.88, per 1 SD in WHR (0.10 unit)) **(Table 1, Figure 2, Supplementary file 2 – figure S2-S3)**. Results were consistent when analyses were stratified by subsite **(Table 1)**. WC, another measure of adiposity did show a protective direction of effect (OR IVW = 0.73, 95%CI 0.52–1.02, *p* = 0.07, per 1 SD increase in WC (0.09 unit)), particularly in the oropharyngeal subsite (OR IVW = 0.66, 95%CI 0.43–1.01, *p* = 0.06, per 1 SD increase in WC (0.09 unit)) (**Table 1, Figure 2, Supplementary file 2 – figure S4**).

**Table 1.** Mendelian randomization results of genetically-proxied metabolic traits with risk of oral and oropharyngeal cancer in GAME-ON

|  |  |  |  |  | IVW |  | Weighted median |  | Weighted mode |  | MR-Egger |  |
| --- | --- | --- | --- | --- | --- | --- | --- | --- | --- | --- | --- | --- |
| Exposure | Outcome | Exposure/<br>Outcome<br>source | Outcome<br>N | Number<br>of SNPs | OR (95%CI) | P | OR (95%CI) | P | OR (95%CI) | P | OR (95%CI) | P |
| BMI | Oral and oropharyngeal cancer combined | Pulit *et al.*<br>GWAS/<br>GAME-ON | 6,034 | 272 | 0.89 (0.72, 1.09) | 0.26 | 0.71 (0.50, 1.00) | 0.05 | 0.63 (0.37, 1.04) | 0.07 | 0.66 (0.40, 1.10) | 0.11 |
|  | Oral cancer |  | 2,990 | 272 | 0.92 (0.71, 1.19) | 0.53 | 0.83 (0.55, 1.28) | 0.40 | 0.79 (0.38, 1.62) | 0.52 | 0.75 (0.39, 1.41) | 0.37 |
|  | Oropharyngeal cancer |  | 2,641 | 272 | 0.89 (0.68, 1.15) | 0.36 | 0.75 (0.50, 1.13) | 0.17 | 0.53 (0.27, 1.03) | 0.06 | 0.56 (0.29, 1.07) | 0.08 |
| WC | Oral and oropharyngeal cancer combined | Shungin *et al.*<br>GWAS/<br>GAME-ON | 6,034 | 43 | 0.73 (0.52, 1.02) | 0.07 | 0.64 (0.40, 1.05) | 0.08 | 0.67 (0.36, 1.26) | 0.22 | 0.43 (0.17, 1.08) | 0.08 |
|  | Oral cancer |  | 2,990 | 43 | 0.82 (0.53, 1.26) | 0.36 | 0.66 (0.36, 1.21) | 0.18 | 0.67 (0.32, 1.39) | 0.29 | 0.54 (0.17, 1.76) | 0.31 |
|  | Oropharyngeal cancer |  | 2,641 | 43 | 0.66 (0.43, 1.01) | 0.06 | 0.56 (0.30, 1.05) | 0.07 | 0.37 (0.17, 0.83) | 0.02 | 0.30 (0.09, 0.98) | 0.05 |
| WHR | Oral and oropharyngeal cancer combined | Pulit *et al.*<br>GWAS/<br>GAME-ON | 6,034 | 176 | 0.98 (0.74, 1.29) | 0.88 | 0.98 (0.64, 1.49) | 0.92 | 0.95 (0.45, 2.00) | 0.89 | 1.80 (0.87, 3.71) | 0.11 |
|  | Oral cancer |  | 2,990 | 176 | 1.18 (0.84, 1.65) | 0.35 | 1.00 (0.58, 1.73) | 0.99 | 0.69 (0.29, 1.67) | 0.41 | 2.49 (1.02, 6.12) | 0.05 |
|  | Oropharyngeal cancer |  | 2,641 | 176 | 0.83 (0.59, 1.14) | 0.25 | 0.88 (0.51, 1.50) | 0.63 | 0.93 (0.37, 2.30) | 0.87 | 1.19 (0.50, 2.86) | 0.70 |
| T2D | Oral and oropharyngeal cancer combined | Vujkovic *et al.*<br>GWAS/<br>GAME-ON | 6,034 | 254 | 0.92 (0.84, 1.01) | 0.09 | 0.85 (0.74, 0.97) | 0.02 | 0.84 (0.71, 1.01) | 0.06 | 0.91 (0.77, 1.09) | 0.31 |
|  | Oral cancer |  | 2,990 | 254 | 0.94 (0.84, 1.05) | 0.27 | 0.84 (0.72, 0.99) | 0.04 | 0.82 (0.66, 1.02) | 0.08 | 0.88 (0.71, 1.08) | 0.22 |
|  | Oropharyngeal cancer |  | 2,641 | 254 | 0.94 (0.84, 1.05) | 0.27 | 0.89 (0.73, 1.10) | 0.29 | 1.02 (0.80, 1.30) | 0.88 | 1.00 (0.81, 1.24) | 0.99 |
| HbA<sub>1c</sub> | Oral and oropharyngeal cancer combined | Wheeler *et al.*<br>GWAS/ | 6,034 | 37 | 0.56 (0.32, 1.00) | 0.05 | 0.52 (0.23, 1.20) | 0.12 | 0.54 (0.24, 1.21) | 0.14 | 0.37 (0.13, 1.05) | 0.07 |
|  |  | GAME-ON<br>(Lesseur et al.,<br>2016) |  |  |  |  |  |  |  |  |  |  |
|  | Oral cancer |  | 2,990 | 37 | 0.48 (0.24, 0.93) | 0.03 | 0.51 (0.18, 1.41) | 0.19 | 0.44 (0.15, 1.29) | 0.14 | 0.30 (0.09, 1.03) | 0.06 |
|  | Oropharyngeal cancer |  | 2,641 | 37 | 0.66 (0.31, 1.40) | 0.28 | 0.49 (0.15, 1.57) | 0.23 | 0.57 (0.18, 1.85) | 0.35 | 0.43 (0.11, 1.68) | 0.23 |
| FG | Oral and oropharyngeal<br>cancer combined | Lagou *et al.*<br>GWAS/<br>GAME-ON<br>(Lesseur et al.,<br>2016) | 6,034 | 28 | 1.06 (0.68, 1.66) | 0.79 | 1.20 (0.62, 2.30) | 0.59 | 1.13 (0.60, 2.12) | 0.71 | 1.11 (0.48, 2.56) | 0.80 |
|  | Oral cancer |  | 2,990 | 28 | 1.05 (0.58, 1.92) | 0.87 | 1.15 (0.48, 2.72) | 0.75 | 0.99 (0.44, 2.23) | 0.99 | 1.25 (0.39, 4.01) | 0.70 |
|  | Oropharyngeal cancer |  | 2,641 | 28 | 1.39 (0.77, 2.51) | 0.28 | 1.24 (0.51, 3.03) | 0.63 | 1.36 (0.59, 3.18) | 0.48 | 1.38 (0.45, 4.18) | 0.58 |
| FI | Oral and oropharyngeal<br>cancer combined | Lagou *et al.*<br>GWAS/<br>GAME-ON | 6,034 | 17 | 0.81 (0.23, 2.89) | 0.75 | 0.75 (0.20, 2.87) | 0.68 | 0.60 (0.03, 10.79) | 0.74 | 0.11 (0.001, 22.47) | 0.43 |
|  | Oral cancer |  | 2,990 | 17 | 0.96 (0.22, 4.16) | 0.96 | 0.46 (0.08, 2.47) | 0.37 | 0.45 (0.01, 19.02) | 0.68 | 0.21 (0.0004, 107.21) | 0.63 |
|  | Oropharyngeal cancer |  | 2,641 | 17 | 0.68 (0.16, 2.87) | 0.59 | 0.66 (0.12, 3.67) | 0.63 | 0.48 (0.05, 4.99) | 0.55 | 0.09 (0.0002, 40.04) | 0.45 |
| SBP | Oral and oropharyngeal<br>cancer combined | Evangelou *et al.*<br>GWAS/<br>GAME-ON<br>(Lesseur et al.,<br>2016) | 6,034 | 83 | 1.00 (0.97, 1.03) | 0.89 | 0.99 (0.94, 1.03) | 0.55 | 0.98 (0.88, 1.09) | 0.66 | 1.06 (0.92, 1.23) | 0.39 |
|  | Oral cancer |  | 2,990 | 83 | 1.01 (0.96, 1.06) | 0.74 | 0.99 (0.93, 1.04) | 0.65 | 0.95 (0.84, 1.08) | 0.48 | 1.09 (0.90, 1.33) | 0.37 |
|  | Oropharyngeal cancer |  | 2,641 | 83 | 0.99 (0.95, 1.03) | 0.65 | 0.99 (0.94, 1.05) | 0.77 | 1.00 (0.88, 1.13) | 0.94 | 1.03 (0.87, 1.23) | 0.71 |
| DBP | Oral and oropharyngeal<br>cancer combined | Evangelou *et al.*<br>GWAS/<br>GAME-ON | 6,034 | 64 | 0.93 (0.87, 1.00) | 0.05 | 0.94 (0.86, 1.04) | 0.22 | 1.10 (0.88, 1.38) | 0.42 | 0.99 (0.80, 1.24) | 0.95 |
|  | Oral cancer |  | 2,990 | 64 | 0.95 (0.87, 1.04) | 0.26 | 0.96 (0.86, 1.07) | 0.45 | 1.17 (0.88, 1.56) | 0.28 | 0.97 (0.74, 1.27) | 0.81 |
|  | Oropharyngeal cancer |  | 2,641 | 64 | 0.92 (0.84, 1.00) | 0.05 | 0.94 (0.84, 1.05) | 0.29 | 1.10 (0.86, 1.41) | 0.45 | 1.00 (0.75, 1.30) | 0.93 |

Abbreviations: IVW, inverse variance weighted; OR, odds ratio; CI, confidence intervals; P, *p*-value; BMI, body mass index; WC, waist circumference; WHR, waist-hip ratio; T2D, type 2 diabetes mellitus; FG, fasting glucose; FI, fasting insulin; HbA<sub>1c</sub>, glycated haemoglobin; SBP, systolic blood pressure; DBP, diastolic blood pressure. OR are expressed per 1 standard deviation (SD) increase in genetically predicted BMI (4.81 kg/m<sup>2</sup>), WC (0.09 unit), WHR (0.10 unit), T2D (1-log unit higher odds of T2D), FG (1-log unit increase in mmol/L fasting glucose), FI (1-log unit increase in mmol/L fasting insulin), HbA<sub>1c</sub> (1-log-unit % higher glycated haemoglobin), SBP (1 unit mmHg increase) and DBP (1 unit mmHg increase).

**Figure 2.**
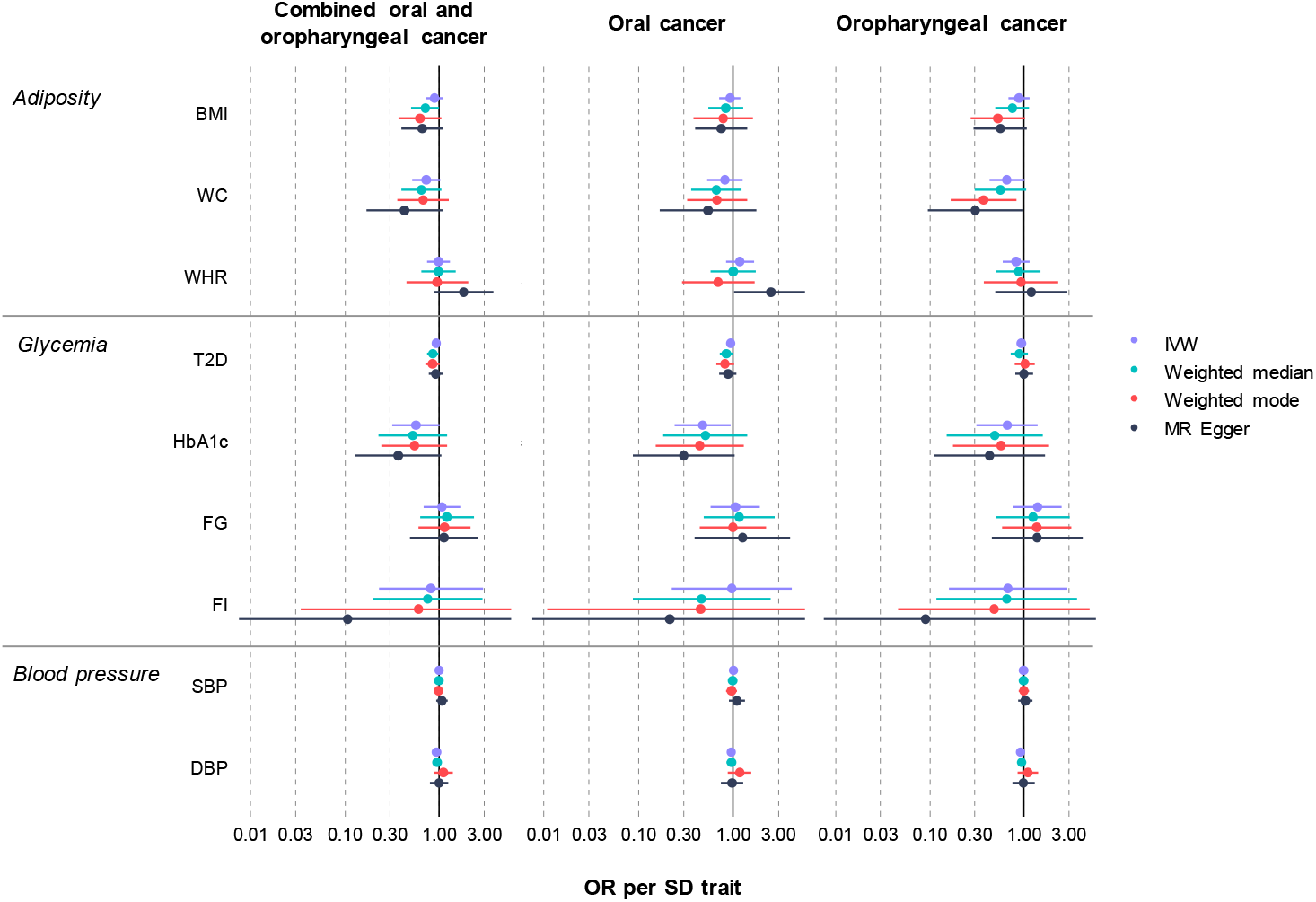
Mendelian randomization results of genetically-proxied metabolic disorders with risk of oral and oropharyngeal cancer including sensitivity analyses in GAME-ON Abbreviations: IVW, inverse variance weighted; OR, odds ratio with 95% confidence intervals; BMI, body mass index; WC, waist circumference; WHR, waist-hip ratio; T2D, type 2 diabetes mellitus; FG, fasting glucose; FI, fasting insulin; HbA1c, glycated haemoglobin; SBP, systolic blood pressure; DBP, diastolic blood pressure. OR are expressed per 1 standard deviation (SD) increase in genetically predicted BMI (4.81 kg/m2), WC (0.09 unit), WHR (0.10 unit), T2D (1-log unit higher odds of T2D), FG (1-log unit increase in mmol/L fasting glucose), FI (1-log unit increase in mmol/L fasting insulin), HbA1c (1-log-unit % higher glycated haemoglobin), SBP (1 unit mmHg increase) and DBP (1 unit mmHg increase).

### Estimated effect of glycaemic traits on oral and oropharyngeal cancer risk

There was limited evidence for an effect of genetically-proxied T2D on combined oral and oropharyngeal cancer (OR IVW = 0.92, 95%CI 0.84–1.01, *p* = 0.09, per 1-log unit higher odds of T2D (**Table 1, Figure 2, Supplementary file 2 – figure S5**). Traits related to diabetes, including HbA_1c_ resulted in a weak protective effect on combined oral and oropharyngeal cancer risk (OR IVW = 0.56, 95%CI 0.32–1.00, *p* = 0.05, per 1-log-unit % higher HbA_1c_), which remained only in the oral subsite (OR IVW = 0.48, 95%CI 0.24–0.93, *p* = 0.03, per 1-log-unit % higher HbA_1c_) following stratification **(Table 1, Figure 2, Supplementary file 2 – figure S6)**. Conversely, there was limited evidence of an effect for FG (OR IVW = 1.06, 95%CI 0.68–1.66, *p* = 0.79, per 1-log unit increase in mmol/L fasting glucose) (Table 1, Figure 2, Supplementary file 2 – figure S7) or FI (OR IVW = 0.81, 95%CI 0.23–2.89, *p* = 0.75, per 1-log unit increase in mmol/L fasting insulin) on combined oral and oropharyngeal cancer risk (**Table 1, Figure 2, Supplementary file 2 – figure S8**)

### Estimated effect of increased blood pressure oral and oropharyngeal cancer risk

Finally, there was limited evidence for an effect of SBP on risk of combined oral and oropharyngeal cancer (OR IVW = 1.00, 95%CI 0.97–1.03, *p* = 0.89, per 1 unit mmHg increase in systolic blood pressure) (**Table 1, Figure 2, Supplementary file 2 – figure S9**), which did not change when stratified by subsite. However, there was some weak evidence for a protective effect of DBP on risk of combined oral and oropharyngeal cancer (OR IVW = 0.93, 95%CI 0.87–1.00, *p* = 0.05, per 1 unit mmHg increase in diastolic blood pressure) (**Table 1, Figure 2, Supplementary file 2 – figure S10**).

### Sensitivity analyses

We conducted MR Egger, weighted median, and weighted mode analyses in addition to IVW (**Table 1, Figure 2**). The results of these analyses generally followed the same pattern as the IVW results reported above, however, there were a number of exceptions. The results for HbA1c were not robust to sensitivity testing (p > 0.05 across methods) (Table 1, Figure 2). In the analysis of T2D on combined oral and oropharyngeal cancer, the weighted median result provided evidence for a weak protective effect (OR weighted median 0.85, 95%CI 0.74–0.97, *p* = 0.02). This effect appeared mainly in the oral subsite (OR weighted median 0.84, 95%CI 0.72–0.99, *p* = 0.04). Furthermore, in the analysis of WC on oropharyngeal cancer risk, the weighted mode supported IVW result, providing evidence of a protective effect (OR weighted mode 0.37, 95%CI 0.17–0.83, *p* = 0.02) (**Table 1, Figure 2**).

There was clear evidence of heterogeneity in the SNP effect estimates OR IVW and MR Egger regression for WHR (Q IVW = 213.04, *p* = 0.03; Q MR Egger = 209.24, *p* = 0.04), T2D (Q IVW = 328.24, p < 0.01; Q MR Egger = 328.21, *p* < 0.01), FI (Q IVW = 32.87, *p* < 0.01; Q MR Egger = 31.63, *p* < 0.01) and DBP (Q IVW = 95.82 p < 0.01; Q MR Egger = 95.22, *p* < 0.01) (**Supplementary file 2 – table S2**). MR-Egger intercepts were not strongly indicative of directional pleiotropy (**Supplementary file 2 – table S3**), but there were outliers present on visual inspection of scatter plots (**Supplementary file 2 – figures S11-19**). MR-PRESSO identified 19 outliers for BMI, 2 outliers for WC, 12 outliers for WHR, 23 outliers for T2D, 4 outliers for HbA_1c_, 1 outlier for FG, 3 outliers for FI, 5 outliers for SBP, 7 outliers for DBP (**Supplementary file 2 – tables S4-5**). When correcting for these outliers, this yielded effects consistent with the primary IVW analysis except for adiposity and T2D instruments, which demonstrated a protective effect on combined oral and oropharyngeal cancer risk when outliers were excluded: BMI (OR IVW = 0.77, 95%CI 0.62–0.94, *p* = 0.01, per 1 SD in BMI (4.81 kg/m^2^)); WC (OR IVW = 0.65, 95%CI 0.47–0.89, *p* = 0.01, per 1 SD in WC (0.09 unit)), and T2D (OR IVW = 0.91, 0.84–0.99, *p* = 0.03, per 1-log unit higher odds of T2D)

(**Supplementary file 2 – table S6**). Where there was evidence of violation of the NOME assumption for WC, FI, SBP and DBP (i.e., I^2^ statistic <0.90) (Supplementary file 2 – table S7), MR-Egger was performed with SIMEX correction. SIMEX effects were consistent with the null, except for SBP where an increased risk effect on combined oral and oropharyngeal cancer was found (OR IVW = 1.15, 95%CI 1.05–1.26, *p* < 0.01, per 1 unit mmHg increase in diastolic blood pressure) (**Supplementary file 2 – table S8**).

### Evaluating instrument-risk factor effects

Where there was some evidence for an effect of BMI, WC, WHR, T2D, HbA_1c_ and DBP on oral and oropharyngeal cancer, we carried out further MR analysis to determine causal effects of these metabolic instruments on established risk HNC risk factors. Obesity-related measures showed a strong causal effect on the risk of smoking initiation: BMI (Beta IVW 0.21 (standard error (SE) 0.03), *p* < 0.001, per 1 SD increase in BMI (4.81 kg/m2)), WC (Beta IVW 0.21 (SE 0.05), *p* < 0.001, per 1 SD increase in WC (0.09 unit)) and WHR (Beta IVW 0.18 (SE 0.03), *p* < 0.001, per 1 SD increase in WHR (0.10 unit)) (**Supplementary file 2 – table S9**). There was weaker evidence for an effect of BMI, WC, and genetic liability to T2D on consumption of alcoholic drinks per week: BMI (Beta IVW -0.04 (SE 0.01), *p* < 0.01, per 1 SD increase in BMI (4.81 kg/m2)), WC (Beta IVW -0.09 (SE 0.02), p < 0.001, per 1 SD increase in WC (0.09 unit)) and T2D (Beta IVW -0.02 (SE 0.01), *p* < 0.001, per 1-log unit higher odds of T2D). BMI (Beta IVW 0.04 (SE 0.01), *p* <0.001, per 1 SD increase in BMI (4.81 kg/m2)) and WHR (Beta IVW 0.04 (SE 0.02), *p* = 0.02, per 1 SD increase in WHR (0.10 unit)) were also estimated to increase general risk tolerance. Similarly, increased BMI or WHR and genetic liability to T2D were estimated to increase educational attainment (years of schooling): BMI (Beta IVW -0.16 (SE 0.02), *p* < 0.001, per 1 SD increase in BMI (4.81 kg/m2)), WHR (Beta IVW -0.11 (SE 0.02), *p* < 0.001, per 1 SD increase in WHR (0.10 unit)), and T2D (Beta IVW -0.02 (SE 0.01), *p* < 0.01, per 1-log unit higher odds of T2D). However, there was strong evidence of both heterogeneity (**Supplementary file 2 – table S10**) and genetic pleiotropy (**Supplementary file 2 – table S11**) across most instrument-risk factor effects. With the exception of alcohol drinks per week, the estimated instrument-risk factor effects remained unchanged following the removal of outlier SNPs detected by MR-PRESSO (**Supplementary file 2 – table S12**).

## Discussion

In this MR study we found limited evidence to support a causal role of genetically-predicted metabolic traits in oral and oropharyngeal cancer, suggesting the risk may have been previously overestimated in observational studies. Where weak evidence for an effect was found (i.e., a protective effect of HbA_1c_), these results were not robust to sensitivity analysis, including outlier correction. There was also strong evidence for instrument-risk factor effects, suggesting smoking may be a mediator between obesity and HNC.

There are several biological mechanisms linking metabolic traits and cancer, but these have not been well explored in HNC (Gatenby & Gillies, 2004; Grimberg, 2003; Tseng, Lin, Lin, & Weng, 2014). Dysregulated metabolism is likely linked to the probability a cancer develops and progresses, given that tumours must adapt to satisfy the bioenergetic and biosynthetic demands of chronic cell proliferation via metabolic reprogramming, enhancing or suppressing the activity of metabolic pathways relative to that in benign tissue (DeNicola & Cantley, 2015). In the largest pooled analysis of 17 case-control studies, BMI was associated with a higher risk of overall HNC, but when stratified by subsite the effect was mainly in the larynx (HR 1.42, 95%CI 1.19–1.70 per 5⍰kg/m^2^, *p* < 0.001) (Gaudet et al., 2015). Laryngeal cancer was not included in our study given that GWAS summary data were not available for this subsite and future analysis of this region is therefore warranted. BMI effects on both the oral (HR 1.10, 95%CI 0.97–1.25, *p* = 0.14) and oropharyngeal cancer (HR 0.98, 95%CI 0.84–1.14, *p* = 0.77) subsites were consistent with the effects found in our study (oral cancer OR 0.92, 95%CI 0.71–1.19, *p* = 0.53; oropharyngeal cancer OR 0.89, 0.68–1.15, *p* = 0.36) (Gaudet et al., 2015). Conversely, the same pooled analysis found an increased risk for both WC (HR 1.09, 95%CI 1.03–1.16, *p* = 0.006) and WHR (HR 1.17, 95%CI 1.02–1.34, *p* = 0.02), mainly in the oral subsite which were not replicated in our MR analysis. Varying patterns of results for these anthropometric measures have been found when stratifying by smoking status within observational studies (Gaudet et al., 2015). The relationship between obesity and HNC is complex. There appears to be a positive association between low BMI (<18.5⍰kg/m^2^) and HNC risk, and a protective effect of BMI on HNC risk in current smokers but conversely, a higher risk in never smokers (Gaudet et al., 2015). This suggests smoking is a confounder, both as an established risk factor for HNC and in its correlation with weight, with nicotine affecting metabolic energy expenditure, leading to reduced calorie absorption and appetite suppression (Williamson et al., 1991). Instrument-risk factor effect estimates from this study suggest smoking is also a mediator, through which metabolic traits such as BMI influence HNC risk.

Despite metabolic syndrome (including hypertension, central obesity, elevated triglyceride, low HDL-C and insulin resistance) being strongly associated with common cancers such as colorectal and breast (Esposito, Chiodini, Colao, Lenzi, & Giugliano, 2012), this does not appear to be the case in head and neck cancer. A recent prospective study of 474,929 participants from UK Biobank investigating the effect of metabolic syndrome suggested those with the condition had no increased HNC risk (HR 1.05, 95%CI 0.90–1.22, *p* = 0.560) (Jiang et al., 2021). No definitive causal effects were detected for individual components of metabolic syndrome components either, supporting our MR results. While another large meta-analysis found individuals with T2D have an elevated risk of oral cancer (Gong et al., 2015), other more recent studies have found this effect to be mostly in laryngeal subsite (HR 1.25, 95%CI 1.12–1.40) which again we could not investigate in this study (H.-B. Kim et al., 2021). Hypertension is the most consistently reported metabolic trait to have an observational association with HNC risk across the subsites (Christakoudi et al., 2020; H.-B. Kim et al., 2021; S.-Y. Kim et al., 2019; Seo et al., 2020; Stocks et al., 2012). We did not identify a clear effect of either SBP or DBP on oral or oropharyngeal cancer using MR, again suggesting the possibility of residual confounding in observational studies.

MR was employed in this study in an attempt to overcome the drawbacks of conventional epidemiological studies. However, there are a number of limitations with using this approach and if MR assumptions are violated, this too can generate spurious conclusions. While there was no evidence of weak instrument bias (F statistics > 10), there was heterogeneity present in at least four of the instruments (WHR, T2D, FI and DBP). This is expected to some extent, given that we are instrumenting multiple biological pathways that contribute to complex metabolic phenotypes. The use of multiple related instruments for each metabolic trait may, however, provide some additional confidence in the overall findings. Given the low percentage of variation explained (*R*^*2*^) for some instruments, as well as the relatively small number of oral and oropharyngeal cancer cases, power to detect an effect may have been an issue in some of our analyses.

As with observational studies, there may be issues of measurement error or misclassification in genetic epidemiology, given BMI is simply a function of mass and height and does not specifically measure adiposity. However, BMI has been shown to be an acceptable proxy when used in large samples sizes, correlating with both total body fat (Browning et al., 2011) and total abdominal adipose tissue (Ross, Léger, Morris, de Guise, & Guardo, 1992), which is thought to present a greater health risk than fat deposited elsewhere. Furthermore, we used a range of adiposity measures including WC and WHR, which may be better proxies of abdominal adiposity, compared to BMI (C. M. Lee, Huxley, Wildman, & Woodward, 2008). SNPs used to proxy these metabolic traits, particularly obesity-related measures BMI, WC and WHR were strongly associated with smoking. Given the heterogeneity of these complex metabolic traits, future work could further examine their pathway specific effects (Udler et al., 2018).

## Conclusion

Overall, there was limited evidence for an effect of genetically-proxied metabolic traits on oral and oropharyngeal cancer risk. These findings suggest metabolic traits may not be effective modifiable risk factors to prioritise as part of future prevention strategies in head and neck cancer. The effect of metabolic traits on the risk of this disease may have been overestimated in previous observational studies, but these cannot be directly compared given the differences in methodological approaches and the interpretation of estimates. Smoking appears to act as a mediator in the relationship between obesity and HNC. Although there is no clear evidence that changing body mass will reduce of increase the risk of HNC directly, dental and medical teams should be aware of the risk of smoking in those who are overweight and therefore the greater risk of cancer when providing smoking cessation and appropriate weight loss advice.

## Supporting information

STROBE MR checklist

## Data Availability

Summary-level analysis was conducted using publicly available GWAS data as cited. Full summary statistics for the GAME-ON outcome data GWAS can be accessed via dbGAP (OncoArray: Oral and Pharynx Cancer; study accession number: phs001202.v1.p1, August 2017) at: https://www.ncbi.nlm.nih.gov/projects/gap/cgi-bin/study.cgi?study_id=phs001202.v1.p1) (Lesseur et al., 2016). This data is also available via the IEU OpenGWAS project (https://gwas.mrcieu.ac.uk/).
All exposure data used in this study is publicly available from the relevant studies as described below. Data for BMI, WC and WHR GWAS was downloaded from the Genetic Investigation of ANthropometric Traits (GIANT) consortium
https://portals.broadinstitute.org/collaboration/giant/index.php/GIANT_consortium_data_files (Pulit et al., 2019; Shungin et al., 2015) and UK Biobank (http://www.ukbiobank.ac.uk). T2D data was downloaded from the DIAMANTE (DIAbetes Meta-ANalysis of Trans-Ethnic association studies) consortium from: https://kp4cd.org/node/169 (Vujkovic et al., 2020). Data for FG, FI and HbA1c, were obtained from GWAS published by the MAGIC (Meta-Analyses of Glucose and Insulin-Related Traits) Consortium, available for download from: https://magicinvestigators.org/downloads/ (Lagou et al., 2021). Finally, data for SBP and DBP were extracted from a GWAS meta-analysis of participants in UK Biobank (http://www.ukbiobank.ac.uk) and the International Consortium of Blood Pressure Genome Wide Association Studies (ICBP), available via dbGAP (International Consortium for Blood Pressure (ICBP), study accession number: phs000585.v2.p1, October 2016) at https://www.ncbi.nlm.nih.gov/projects/gap/cgi-bin/study.cgi?study_id=phs000585.v2.p1 (Evangelou et al., 2018).
Instrument-risk factor analysis outcome summary-level data were derived from the GWAS & Sequencing Consortium of Alcohol and Nicotine use (GSCAN) and UK Biobank and UK Biobank (http://www.ukbiobank.ac.uk) for alcoholic drinks per week https://conservancy.umn.edu/handle/11299/201564 (Liu et al., 2019) and the comprehensive smoking index (Wootton et al., 2019). Data for risk tolerance and educational attainment were taken from Social Science Genetic Association Consortium (SSGAC) data available from http://www.thessgac.org/data (Karlsson Linner et al., 2019; J. Lee et al., 2018). MR analyses were conducted using the TwoSampleMR package in R (version 3.5.3). A copy of the code and all data files used in this study are available at GitHub (https://github.com/MGormley12/metabolic_trait_hnc_mr.git).

https://github.com/MGormley12/metabolic_trait_hnc_mr.git

https://portals.broadinstitute.org/collaboration/giant/index.php/GIANT_consortium_data_files

https://kp4cd.org/node/169

https://magicinvestigators.org/downloads/

http://www.ukbiobank.ac.uk)

https://www.ncbi.nlm.nih.gov/projects/gap/cgi-bin/study.cgi?study_id=phs000585.v2.p1

## Ethics approval and consent to participate

All studies included as part of the GAME-ON network obtained approval and consent from their respective institutions.

## Consent for publication

Not applicable.

## Availability of data and materials

Summary-level analysis was conducted using publicly available GWAS data as cited. Full summary statistics for the GAME-ON outcome data GWAS can be accessed via dbGAP (OncoArray: Oral and Pharynx Cancer; study accession number: phs001202.v1.p1, August 2017) at: https://www.ncbi.nlm.nih.gov/projects/gap/cgi-bin/study.cgi?study_id=phs001202.v1.p1) (Lesseur et al., 2016). This data is also available via the IEU OpenGWAS project (https://gwas.mrcieu.ac.uk/).

All exposure data used in this study is publicly available from the relevant studies as described below. Data for BMI, WC and WHR GWAS was downloaded from the Genetic Investigation of ANthropometric Traits (GIANT) consortium https://portals.broadinstitute.org/collaboration/giant/index.php/GIANT_consortium_data_files (Pulit et al., 2019; Shungin et al., 2015) and UK Biobank (http://www.ukbiobank.ac.uk). T2D data was downloaded from the DIAMANTE (DIAbetes Meta-ANalysis of Trans-Ethnic association studies) consortium from: https://kp4cd.org/node/169 (Vujkovic et al., 2020). Data for FG, FI and HbA1c, were obtained from GWAS published by the MAGIC (Meta-Analyses of Glucose and Insulin-Related Traits) Consortium, available for download from: https://magicinvestigators.org/downloads/ (Lagou et al., 2021). Finally, data for SBP and DBP were extracted from a GWAS meta-analysis of participants in UK Biobank http://www.ukbiobank.ac.uk) and the International Consortium of Blood Pressure Genome Wide Association Studies (ICBP), available via dbGAP (International Consortium for Blood Pressure (ICBP), study accession number: phs000585.v2.p1, October 2016) at https://www.ncbi.nlm.nih.gov/projects/gap/cgi-bin/study.cgi?study_id=phs000585.v2.p1 (Evangelou et al., 2018).

Instrument-risk factor analysis outcome summary-level data were derived from the GWAS & Sequencing Consortium of Alcohol and Nicotine use (GSCAN) and UK Biobank and UK Biobank (http://www.ukbiobank.ac.uk) for alcoholic drinks per week https://conservancy.umn.edu/handle/11299/201564 (Liu et al., 2019) and the comprehensive smoking index (Wootton et al., 2019). Data for risk tolerance and educational attainment were taken from Social Science Genetic Association Consortium (SSGAC) data available from http://www.thessgac.org/data (Karlsson Linner et al., 2019; J. Lee et al., 2018). MR analyses were conducted using the “TwoSampleMR” package in R (version 3.5.3). A copy of the code and all data files used in this study are available at GitHub (https://github.com/MGormley12/metabolic_trait_hnc_mr.git).

