## Supplementary file 2 for "Evaluating the effect of metabolic traits on oral and oropharyngeal cancer risk using Mendelian randomization"

**Table S1.** Assessing weak instrument bias (F-statistic) and proportion of variance in the phenotype (*R*^2^) explained by metabolic phenotype instruments

|  | R^2^ | F-statistic |
| --- | --- | --- |
| **BMI** | 0.040 (4.0%) | 89.5 |
| **WC** | 0.012 (1.2%) | 60.1 |
| **WHR** | 0.023 (2.3%) | 76.1 |
| **T2D** | 0.022 (2.2%) | 113.7 |
| **HbA_1c_** | 0.018 (1.8%) | 70.4 |
| **FG** | 0.030 (3%) | 133.6 |
| **FI** | 0.005 (0.5%) | 33.3 |
| **SBP** | 0.008 (0.6%) | 56.9 |
| **DBP** | 0.006 (0.6%) | 56.5 |

Abbreviations: BMI, body mass index; WC, waist circumference; WHR, waist-hip ratio; T2D, type 2 diabetes mellitus; HbA_1c_, glycated haemoglobin; FG, fasting glucose; FI, fasting insulin; SBP, systolic blood pressure; DBP, diastolic blood pressure.

**Table S2.** Assessing heterogeneity of single nucleotide polymorphism effect estimates in inverse-variance weighted (IVW) and MR Egger regression for metabolic disorder analysis.

| **Exposure** | **Exposure dataset** | **Q IVW** | **df** | **P** | **Q MR Egger** | **df** | **P** |
| --- | --- | --- | --- | --- | --- | --- | --- |
| BMI | Pulit *et al.* GWAS ^1^ | 311.47 | 271 | 0.05 | 309.67 | 270 | 0.05 |
| WC | Pulit *et al.* GWAS ^1^ | 50.35 | 42 | 0.18 | 48.63 | 41 | 0.19 |
| WHR | Shungin *et al.* GWAS ^2^ | 213.04 | 175 | 0.03 | 209.24 | 174 | 0.04 |
| T2D | Vujkovic *et al.* GWAS ^3^ | 328.24 | 253 | <0.01 | 328.21 | 252 | <0.01 |
| HbA_1c_ | Wheeler *et al.* GWAS ^4^ | 40.40 | 36 | 0.28 | 39.35 | 35 | 0.28 |
| FG | Lagou *et al.* GWAS ^5^ | 24.00 | 27 | 0.63 | 23.98 | 26 | 0.58 |
| FI | Lagou *et al.* GWAS ^5^ | 32.87 | 16 | <0.01 | 31.63 | 15 | <0.01 |
| SBP | Evangelou *et al.* GWAS ^6^ | 95.79 | 82 | 0.15 | 94.81 | 82 | 0.14 |
| DBP | Evangelou *et al.* GWAS ^6^ | 95.82 | 63 | <0.01 | 95.22 | 62 | <0.01 |

Abbreviations: Q, Q-statistic; df, degrees of freedom; P, p-value; BMI, body mass index; WC, waist circumference; WHR, waist-hip ratio; T2D, type 2 diabetes mellitus; HbA_1c_, glycated haemoglobin; FG, fasting glucose; FI, fasting insulin; SBP, systolic blood pressure; DBP, diastolic blood pressure.

**Table S3.** Assessing directional pleiotropy through MR Egger intercept for metabolic disorder analysis.

| **Exposure** | **Exposure dataset** | **N SNPs** | **Estimate** | **SE** | **P** |
| --- | --- | --- | --- | --- | --- |
| BMI | Pulit *et al.* GWAS ^1^ | 272 | 0.006 | 0.005 | 0.21 |
| WC | Pulit *et al.* GWAS ^1^ | 43 | 0.016 | 0.013 | 0.24 |
| WHR | Shungin *et al.* GWAS ^2^ | 176 | -0.011 | 0.006 | 0.08 |
| T2D | Vujkovic *et al.* GWAS ^3^ | 254 | 0.001 | 0.004 | 0.88 |
| HbA_1c_ | Wheeler *et al.* GWAS ^4^ | 37 | 0.009 | 0.010 | 0.34 |
| FG | Lagou *et al.* GWAS ^5^ | 28 | -0.001 | 0.011 | 0.90 |
| FI | Lagou *et al.* GWAS ^5^ | 17 | 0.032 | 0.042 | 0.46 |
| SBP | Evangelou *et al.* GWAS ^6^ | 83 | -0.014 | 0.016 | 0.36 |
| DBP | Evangelou *et al.* GWAS ^6^ | 64 | -0.010 | 0.015 | 0.54 |

Abbreviations: SE, standard error; P, p-value; BMI, body mass index; WC, waist circumference; WHR, waist-hip ratio; T2D, type 2 diabetes mellitus; HbA_1c_, glycated haemoglobin; FG, fasting glucose; FI, fasting insulin; SBP, systolic blood pressure; DBP, diastolic blood pressure.

**Table S4.** MR-PRESSO outliers detected results in the analysis of metabolic disorders on combined oral and oropharyngeal cancer risk.

| **Exposure** | **SNP** | **Q statistic** | **P value** | **Q sum** | **Q difference** |
| --- | --- | --- | --- | --- | --- |
| BMI | rs1928295 | 9.75 | <0.001 | 9.75 | -301.70 |
|  | rs17201143 | 9.26 | <0.001 | 19.01 | -292.44 |
|  | rs1320251 | 8.17 | <0.001 | 27.18 | -284.27 |
|  | rs13107325 | 6.38 | 0.01 | 33.56 | -277.89 |
|  | rs3808477 | 6.12 | 0.01 | 39.68 | -271.77 |
|  | rs12140153 | 5.82 | 0.02 | 45.50 | -265.94 |
|  | rs194809 | 5.80 | 0.02 | 51.30 | -260.15 |
|  | rs11655587 | 5.65 | 0.02 | 56.96 | -254.49 |
|  | rs12072739 | 5.57 | 0.02 | 62.53 | -248.92 |
|  | rs12321904 | 5.11 | 0.02 | 67.64 | -243.81 |
|  | rs5215 | 4.99 | 0.03 | 72.63 | -238.82 |
|  | rs2228213 | 4.85 | 0.03 | 77.48 | -233.97 |
|  | rs12282785 | 4.73 | 0.03 | 82.21 | -229.24 |
|  | rs10929925 | 4.34 | 0.04 | 86.55 | -224.90 |
|  | rs2862996 | 4.28 | 0.04 | 90.82 | -220.63 |
|  | rs13021737 | 4.02 | 0.04 | 94.85 | -216.60 |
|  | rs561136 | 4.01 | 0.05 | 98.86 | -212.59 |
|  | rs11047132 | 3.95 | 0.05 | 102.81 | -208.64 |
|  | rs7713317 | 3.90 | 0.05 | 106.71 | -204.74 |
| WC | rs1928295 | 10.74 | <0.001 | 10.74 | -39.53 |
|  | rs806794 | 7.42 | 0.01 | 18.16 | -32.11 |
| WHR | rs12777288 | 12.17 | <0.001 | 12.17 | -200.87 |
|  | rs7213608 | 8.21 | <0.001 | 20.38 | -192.66 |
|  | rs4395620 | 7.34 | 0.01 | 27.72 | -185.32 |
|  | rs2167750 | 6.78 | 0.01 | 34.51 | -178.53 |
|  | rs55747707 | 6.25 | 0.01 | 40.75 | -172.29 |
|  | rs12140153 | 6.16 | 0.01 | 46.92 | -166.13 |
|  | rs12608504 | 5.59 | 0.02 | 52.51 | -160.53 |
|  | rs76699125 | 5.59 | 0.02 | 58.10 | -154.95 |
|  | rs1787013 | 5.05 | 0.02 | 63.14 | -149.90 |
|  | rs6688233 | 5.00 | 0.03 | 68.14 | -144.90 |
|  | rs861029 | 4.93 | 0.03 | 73.07 | -139.98 |
|  | rs17738166 | 4.34 | 0.04 | 77.41 | -135.63 |
| T2D | rs6011155 | 17.92 | <0.01 | 17.92 | -310.28 |
|  | rs12789028 | 8.23 | <0.01 | 26.15 | -302.05 |
|  | rs4833687 | 7.10 | 0.01 | 33.24 | -294.95 |
|  | rs7483027 | 6.54 | 0.01 | 39.78 | -288.42 |
|  | rs10808671 | 6.49 | 0.01 | 46.27 | -281.93 |
|  | rs2269247 | 6.06 | 0.01 | 52.32 | -275.87 |
|  | rs3753693 | 5.92 | 0.01 | 58.25 | -269.95 |
|  | rs1470560 | 5.88 | 0.02 | 64.13 | -264.07 |
|  | rs11759026 | 5.67 | 0.02 | 69.80 | -258.40 |
|  | rs12918782 | 5.47 | 0.02 | 75.27 | -252.93 |
|  | rs174541 | 5.46 | 0.02 | 80.73 | -247.47 |
|  | rs4671799 | 5.20 | 0.02 | 85.93 | -242.27 |
|  | rs3130931 | 5.11 | 0.02 | 91.04 | -237.16 |
|  | rs7664347 | 4.35 | 0.04 | 95.38 | -232.81 |
|  | rs61817176 | 4.30 | 0.04 | 99.69 | -228.51 |
|  | rs13059382 | 4.25 | 0.04 | 103.94 | -224.26 |
|  | rs429358 | 4.23 | 0.04 | 108.17 | -220.03 |
|  | rs60384372 | 4.21 | 0.04 | 112.38 | -215.82 |
|  | rs3744347 | 4.19 | 0.04 | 116.57 | -211.62 |
|  | rs73121277 | 4.11 | 0.04 | 120.69 | -207.51 |
|  | rs757110 | 3.93 | 0.05 | 124.61 | -203.58 |
|  | rs6976111 | 3.93 | 0.05 | 128.54 | -199.66 |
|  | rs10916780 | 3.92 | 0.05 | 132.46 | -195.74 |
| HbA_1c_ | rs4783565 | 11.33 | <0.001 | 11.33 | -29.01 |
|  | rs10774625 | 4.72 | 0.03 | 16.05 | -24.29 |
|  | rs11248914 | 4.01 | 0.05 | 20.06 | -20.29 |
|  | rs17747324 | 3.96 | 0.05 | 24.01 | -16.33 |
| FG | rs983309 | 4.41 | 0.04 | 4.41 | -19.59 |
| FI | rs1167800 | 6.01 | 0.01 | 6.01 | -26.84 |
|  | rs35767 | 5.12 | 0.02 | 11.14 | -21.71 |
|  | rs3822072 | 5.10 | 0.02 | 16.23 | -16.62 |
| SBP | rs2613765 | 7.93 | <0.001 | 7.93 | -87.86 |
|  | rs10069690 | 7.64 | 0.01 | 15.57 | -80.22 |
|  | rs7187540 | 6.59 | 0.01 | 22.16 | -73.63 |
|  | rs260508 | 4.87 | 0.03 | 27.03 | -68.76 |
|  | rs4651224 | 3.95 | 0.05 | 30.98 | -64.81 |
| DBP | rs360153 | 9.71 | <0.001 | 9.71 | -85.95 |
|  | rs11026586 | 5.61 | 0.02 | 15.32 | -80.34 |
|  | rs1607644 | 5.60 | 0.02 | 20.92 | -74.74 |
|  | rs4411245 | 5.27 | 0.02 | 26.19 | -69.47 |
|  | rs61892344 | 4.36 | 0.04 | 30.55 | -65.11 |
|  | rs954767 | 4.11 | 0.04 | 34.67 | -60.99 |
|  | rs1718845 | 3.94 | 0.05 | 38.61 | -57.05 |

Abbreviations: Q-stat, Cochran’s Q statistic; BMI, body mass index; WC, waist circumference; WHR, waist-hip ratio; T2D, type 2 diabetes mellitus; HbA_1c_, glycated haemoglobin; FG, fasting glucose; FI, fasting insulin; SBP, systolic blood pressure; DBP, diastolic blood pressure.

**Table S5.** MR-PRESSO results for metabolic disorders on combined oral and oropharyngeal cancer.

| **Exposure** | **Outcome** | **Global test RSSobs** | **P-value** | **Outlier test** | **P-value** |
| --- | --- | --- | --- | --- | --- |
| BMI | Oral and oropharyngeal cancer | 314.14 | 0.05 | <0.01 | 1.00 |
| WC | Oral and oropharyngeal cancer | 52.43 | 0.22 | NA | NA |
| WHR | Oral and oropharyngeal cancer | 215.50 | 0.04 | <0.01 | 1.00 |
| T2D | Oral and oropharyngeal cancer | 330.88 | <0.01 | <0.01 | 1.00 |
| HbA_1c_ | Oral and oropharyngeal cancer | 41.66 | 0.33 | NA | NA |
| FG | Oral and oropharyngeal cancer | 25.34 | 0.68 | NA | NA |
| FI | Oral and oropharyngeal cancer | 38.87 | 0.01 | 1.42E-05 | 1.00 |
| SBP | Oral and oropharyngeal cancer | 98.16 | 0.15 | NA | NA |
| DBP | Oral and oropharyngeal cancer | 99.40 | <0.01 | <0.01 | 1.00 |

Abbreviations: SE, standard error; P, p-value; BMI, body mass index; WC, waist circumference; WHR, waist-hip ratio; T2D, type 2 diabetes mellitus; HbA_1c_, glycated haemoglobin; FG, fasting glucose; FI, fasting insulin; SBP, systolic blood pressure; DBP, diastolic blood pressure.

**Table S6.** Outlier corrected results in the analysis of metabolic disorders on combined oral and oropharyngeal cancer risk.

| **Exposure** | **N SNP** | **Method** | **Beta** | **SE** | **OR** | **CIL** | **CIU** | **P value** |
| --- | --- | --- | --- | --- | --- | --- | --- | --- |
| BMI | 253 | IVW | -0.27 | 0.10 | 0.77 | 0.62 | 0.94 | 0.01 |
|  | 253 | MR Egger | -0.39 | 0.26 | 0.68 | 0.41 | 1.13 | 0.13 |
|  | 253 | Weighted median | -0.41 | 0.17 | 0.67 | 0.48 | 0.93 | 0.02 |
|  | 253 | Weighted mode | -0.49 | 0.25 | 0.61 | 0.37 | 1.00 | 0.05 |
| WC | 41 | IVW | -0.43 | 0.16 | 0.65 | 0.47 | 0.89 | 0.01 |
|  | 41 | MR Egger | -0.6 | 0.44 | 0.55 | 0.23 | 1.31 | 0.18 |
|  | 41 | Weighted median | -0.44 | 0.24 | 0.64 | 0.40 | 1.04 | 0.07 |
|  | 41 | Weighted mode | -0.4 | 0.31 | 0.67 | 0.36 | 1.24 | 0.21 |
| WHR | 164 | IVW | -0.13 | 0.13 | 0.87 | 0.68 | 1.13 | 0.30 |
|  | 164 | MR Egger | 0.5 | 0.34 | 1.65 | 0.84 | 3.24 | 0.15 |
|  | 164 | Weighted median | -0.04 | 0.21 | 0.96 | 0.64 | 1.45 | 0.86 |
|  | 164 | Weighted mode | -0.06 | 0.41 | 0.94 | 0.43 | 2.09 | 0.88 |
| T2D | 231 | IVW | -0.09 | 0.04 | 0.91 | 0.84 | 0.99 | 0.03 |
|  | 231 | MR Egger | -0.04 | 0.08 | 0.96 | 0.82 | 1.12 | 0.60 |
|  | 231 | Weighted median | -0.16 | 0.07 | 0.85 | 0.74 | 0.97 | 0.02 |
|  | 231 | Weighted mode | -0.2 | 0.1 | 0.82 | 0.67 | 0.99 | 0.04 |
| HbA_1c_ | 33 | IVW | -0.55 | 0.29 | 0.58 | 0.33 | 1.01 | 0.05 |
|  | 33 | MR Egger | -0.75 | 0.51 | 0.47 | 0.17 | 1.30 | 0.16 |
|  | 33 | Weighted median | -0.65 | 0.43 | 0.52 | 0.23 | 1.22 | 0.13 |
|  | 33 | Weighted mode | -0.63 | 0.45 | 0.53 | 0.22 | 1.29 | 0.17 |
| FG | 27 | IVW | 0.14 | 0.23 | 1.15 | 0.73 | 1.80 | 0.55 |
|  | 27 | MR Egger | 0.18 | 0.43 | 1.19 | 0.52 | 2.75 | 0.68 |
|  | 27 | Weighted median | 0.18 | 0.33 | 1.2 | 0.63 | 2.28 | 0.59 |
|  | 27 | Weighted mode | 0.12 | 0.31 | 1.12 | 0.61 | 2.06 | 0.71 |
| FI | 14 | IVW | -0.97 | 0.51 | 0.38 | 0.14 | 1.02 | 0.06 |
|  | 14 | MR Egger | -0.86 | 2.47 | 0.42 | 0.01 | 53.86 | 0.73 |
|  | 14 | Weighted median | -0.4 | 0.74 | 0.67 | 0.16 | 2.83 | 0.59 |
|  | 14 | Weighted mode | -2.88 | 1.68 | 0.06 | 0.01 | 1.50 | 0.11 |
| SBP | 78 | IVW | -0.01 | 0.02 | 0.99 | 0.96 | 1.02 | 0.46 |
|  | 78 | MR Egger | -0.01 | 0.07 | 0.99 | 0.87 | 1.14 | 0.94 |
|  | 78 | Weighted median | -0.02 | 0.02 | 0.98 | 0.94 | 1.03 | 0.49 |
|  | 78 | Weighted mode | -0.02 | 0.05 | 0.98 | 0.88 | 1.08 | 0.63 |
| DBP | 57 | IVW | -0.06 | 0.03 | 0.94 | 0.89 | 1.00 | 0.06 |
|  | 57 | MR Egger | -0.02 | 0.1 | 0.98 | 0.81 | 1.19 | 0.83 |
|  | 57 | Weighted median | -0.05 | 0.05 | 0.96 | 0.87 | 1.05 | 0.32 |
|  | 57 | Weighted mode | 0.12 | 0.11 | 1.12 | 0.91 | 1.40 | 0.29 |

Abbreviations: SE, standard error; OR, odds ratio; CI, confidence intervals; IVW, inverse variance weighted; BMI, body mass index; WC, waist circumference; WHR, waist-hip ratio; T2D, type 2 diabetes mellitus; HbA_1c_, glycated haemoglobin; FG, fasting glucose; FI, fasting insulin; SBP, systolic blood pressure; DBP, diastolic blood pressure.

**Table S7.** Assessing violation of the NO Measurement Error (NOME) assumption for instruments used in MR-Egger regression.

| **Exposure** | **Exposure dataset** | **I^2^ unweighted** | **I^2^ weighted** |
| --- | --- | --- | --- |
| BMI | Pulit *et al.* GWAS ^1^ | 0.94 | 0.93 |
| WC | Pulit *et al.* GWAS ^1^ | 0.89 | 0.87 |
| WHR | Shungin *et al.* GWAS ^2^ | 0.90 | 0.88 |
| T2D | Vujkovic *et al.* GWAS ^3^ | 0.97 | 0.97 |
| HbA_1c_ | Wheeler *et al.* GWAS ^4^ | 0.94 | 0.92 |
| FG | Lagou *et al.* GWAS ^5^ | 0.98 | 0.98 |
| FI | Lagou *et al.* GWAS ^5^ | 0.55 | 0.37 |
| SBP | Evangelou *et al.* GWAS ^6^ | 0.72 | 0.35 |
| DBP | Evangelou *et al.* GWAS ^6^ | 0.81 | 0.59 |

Abbreviations: I^2^, I-squared statistic; BMI, body mass index; WC, waist circumference; WHR, waist-hip ratio; T2D, type 2 diabetes mellitus; HbA_1c_, glycated haemoglobin; FG, fasting glucose; FI, fasting insulin; SBP, systolic blood pressure; DBP, diastolic blood pressure.

**Table S8.** SIMEX correction MR Egger regression results for where NO Measurement Error (NOME) assumption may have been violated (I^2^ <0.90).

| **Exposure** | **Outcome** | **OR** | **CIL** | **CIU** | **P-value** |
| --- | --- | --- | --- | --- | --- |
| WC | Oral and oropharyngeal cancer | 0.64 | 0.25 | 1.63 | 0.36 |
| FI | Oral and oropharyngeal cancer | 0.01 | 2.10E-05 | 4.35 | 0.16 |
| SBP | Oral and oropharyngeal cancer | 1.15 | 1.05 | 1.26 | <0.01 |
| DBP | Oral and oropharyngeal cancer | 1.07 | 0.91 | 1.25 | 0.43 |

Abbreviations: OR, odds ratio; CI, confidence intervals; WC, waist circumference; FI, fasting insulin; SBP, systolic blood pressure; DBP, diastolic blood pressure.

**Table S9.** Mendelian randomization results evaluating instrument-risk factor effects

|  |  | | | **IVW** | | **Weighted median** | | **Weighted mode** | | **MR-Egger** | |
| --- | --- | --- | --- | --- | --- | --- | --- | --- | --- | --- | --- |
| **Exposure** | **Outcome** | **Exposure/**  **Outcome**  **source** | **N SNPs** | **Beta (SE)** | **P** | **Beta (SE)** | **P** | **Beta (SE)** | **P** | **Beta (SE)** | **P** |
| BMI | Smoking initiation | Pulit *et al.* GWAS ^1^/ Liu et al. GWAS ^7^ | 273 | 0.21 (0.03) | <0.001 | 0.14 (0.03) | <0.001 | 0.08 (0.04) | 0.07 | 0.07 (0.06) | 0.27 |
|  | Alcohol drinks per week | Pulit *et al.* GWAS ^1^/ Liu et al. GWAS ^7^ | 274 | -0.04 (0.01) | 0.01 | -0.05 (0.01) | <0.001 | -0.10 (0.02) | <0.001 | -0.11 (0.03) | <0.001 |
|  | Risk tolerance | Pulit *et al.* GWAS ^1^/ Karlsson Linner et al. GWAS ^8^ | 301 | 0.04 (0.01) | <0.001 | 0.001 (0.02) | 0.92 | -0.06 (0.03) | 0.05 | -0.03 (0.03) | 0.41 |
|  | Educational attainment | Pulit *et al.* GWAS ^1^/ Lee et al. GWAS ^9^ | 299 | -0.16 (0.02) | <0.001 | -0.11 (0.01) | <0.001 | -0.03 (0.03) | 0.47 | 0.01 (0.04) | 0.82 |
| WC | Smoking initiation | Pulit *et al.* GWAS ^1^/ Liu et al. GWAS ^7^ | 43 | 0.21 (0.05) | <0.001 | 0.09 (0.04) | 0.03 | 0.05 (0.04) | 0.25 | -0.13 (0.14) | 0.35 |
|  | Alcohol drinks per week | Pulit *et al.* GWAS ^1^/ Liu et al. GWAS ^7^ | 43 | -0.09 (0.02) | <0.001 | -0.11 (0.02) | <0.001 | -0.12 (0.02) | <0.001 | -0.20 (0.06) | <0.001 |
|  | Risk tolerance | Pulit *et al.* GWAS ^1^/ Karlsson Linner et al. GWAS ^8^ | 45 | 0.01 (0.02) | 0.71 | -0.04 (0.02) | 0.05 | -0.05 (0.03) | 0.08 | -0.07 (0.06) | 0.27 |
|  | Educational attainment | Pulit *et al.* GWAS ^1^/ Lee et al. GWAS ^9^ | 45 | -0.06 (0.03) | 0.06 | -0.06 (0.02) | 0.01 | 0.01 (0.02) | 0.77 | 0.08 (0.08) | 0.34 |
| WHR | Smoking initiation | Shungin *et al.* GWAS ^2^/ Liu et al. GWAS ^7^ | 174 | 0.18 (0.03) | <0.001 | 0.09 (0.03) | <0.001 | 0.07 (0.04) | 0.12 | 0.02 (0.09) | 0.84 |
|  | Alcohol drinks per week | Shungin *et al.* GWAS ^2^/ Liu et al. GWAS ^7^ | 174 | -0.03 (0.02) | 0.11 | -0.02 (0.02) | 0.21 | -0.02 (0.02) | 0.30 | -0.07 (0.04) | 0.11 |
|  | Risk tolerance | Shungin *et al.* GWAS ^2^/ Karlsson Linner et al. GWAS ^8^ | 199 | 0.04 (0.02) | 0.02 | -0.001 (0.02) | 0.96 | -0.04 (0.03) | 0.18 | -0.04 (0.04) | 0.32 |
|  | Educational attainment | Shungin *et al.* GWAS ^2^/ Lee et al. GWAS ^9^ | 196 | -0.11 (0.02) | <0.001 | -0.03 (0.02) | 0.10 | 0.02 (0.02) | 0.40 | 0.08 (0.06) | 0.15 |
| T2D | Smoking initiation | Vujkovic *et al.* GWAS ^3^/ Liu et al. GWAS ^7^ | 255 | 0.02 (0.01) | 0.12 | 0.02 (0.01) | 0.08 | 0.02 (0.01) | 0.08 | -0.01 (0.02) | 0.68 |
|  | Alcohol drinks per week | Vujkovic *et al.* GWAS ^3^/ Liu et al. GWAS ^7^ | 257 | -0.02 (0.01) | <0.001 | -0.02 (0.01) | <0.001 | -0.02 (0.01) | <0.001 | -0.02 (0.01) | 0.11 |
|  | Risk tolerance | Vujkovic *et al.* GWAS ^3^/ Karlsson Linner et al. GWAS ^8^ | 274 | -0.0002 (0.004) | 0.96 | -0.01 (0.01) | 0.38 | -0.01 (0.01) | 0.30 | -0.01 (0.01) | 0.15 |
|  | Educational attainment | Vujkovic *et al.* GWAS ^3^/ Lee et al. GWAS ^9^ | 272 | -0.02 (0.01) | 0.01 | -0.01 (0.01) | 0.11 | -0.0001 (0.004) | 0.97 | 0.02 (0.01) | 0.10 |
| HbA_1c_ | Smoking initiation | Wheeler *et al.* GWAS ^4^/ Liu et al. GWAS ^7^ | 37 | 0.03 (0.06) | 0.64 | 0.001 (0.06) | 0.99 | 0.002 (0.05) | 0.98 | 0.11 (0.10) | 0.30 |
|  | Alcohol drinks per week | Wheeler *et al.* GWAS ^4^/ Liu et al. GWAS ^7^ | 37 | 0.01 (0.03) | 0.69 | 0.05 (0.03) | 0.12 | 0.08 (0.03) | 0.02 | 0.11 (0.05) | 0.04 |
|  | Risk tolerance | Wheeler *et al.* GWAS ^4^/ Karlsson Linner et al. GWAS ^8^ | 40 | -0.001 (0.02) | 0.97 | 0.01 (0.03) | 0.68 | 0.01 (0.03) | 0.69 | 0.003 (0.04) | 0.95 |
|  | Educational attainment | Wheeler *et al.* GWAS ^4^/ Lee et al. GWAS ^9^ | 40 | -0.01 (0.04) | 0.80 | -0.02 (0.03) | 0.56 | 0.04 (0.04) | 0.30 | -0.001 (0.08) | 0.99 |
| DBP | Smoking initiation | Evangelou *et al.* GWAS ^6^/ Liu et al. GWAS ^7^ | 64 | -0.01 (0.01) | 0.24 | -0.0004 (0.01) | 0.95 | 0.01 (0.01) | 0.45 | 0.004 (0.02) | 0.85 |
|  | Alcohol drinks per week | Evangelou *et al.* GWAS ^6^/ Liu et al. GWAS ^7^ | 64 | 0.0004 (0.004) | 0.91 | 0.001 (0.003) | 0.79 | 0.005 (0.01) | 0.44 | -0.003 (0.01) | 0.77 |
|  | Risk tolerance | Evangelou *et al.* GWAS ^6^/ Karlsson Linner et al. GWAS ^8^ | 75 | 0.002 (0.003) | 0.53 | 0.002 (0.003) | 0.53 | 0.004 (0.01) | 0.64 | 0.017 (0.01) | 0.05 |
|  | Educational attainment | Evangelou *et al.* GWAS ^6^/ Lee et al. GWAS ^9^ | 74 | 0.001 (0.004) | 0.75 | -0.001 (0.003) | 0.67 | -0.01 (0.008) | 0.11 | 0.02 (0.01) | 0.06 |

Abbreviations: IVW, inverse variance weighted; OR, odds ratio; CI, confidence intervals; P, p-value; BMI, body mass index; WC, waist circumference; WHR, waist-hip ratio; T2D, type 2 diabetes mellitus;, glycated haemoglobin; DBP, diastolic blood pressure. OR are expressed per 1 standard deviation (SD) increase in genetically predicted BMI (4.81 kg/m2), WC (0.09 unit), WHR (0.10 unit), T2D (1-log unit higher odds of T2D), HbA1c (1-log-unit % higher glycated haemoglobin), and DBP (1 unit mmHg increase). Outcome beta estimates reflect the standard deviation of the phenotype.

**Table S10.** Assessing heterogeneity in Mendelian randomization results evaluating instrument-risk factor effects

| **Exposure** | **Outcome** | **Q IVW** | **df** | **P** | **Q MR Egger** | **df** | **P** |
| --- | --- | --- | --- | --- | --- | --- | --- |
| BMI | Smoking initiation | 1033.81 | 272 | <0.001 | 1011.43 | 271 | <0.001 |
|  | Alcohol drinks per week | 952.47 | 273 | <0.001 | 928.23 | 272 | <0.001 |
|  | Risk tolerance | 831.50 | 300 | <0.001 | 813.38 | 299 | <0.001 |
|  | Educational attainment | 1939.06 | 298 | <0.001 | 1798.21 | 297 | <0.001 |
| WC | Smoking initiation | 271.19 | 42 | <0.001 | 231.18 | 41 | <0.001 |
|  | Alcohol drinks per week | 177.18 | 42 | <0.001 | 163.17 | 41 | <0.001 |
|  | Risk tolerance | 155.99 | 44 | <0.001 | 149.70 | 43 | <0.001 |
|  | Educational attainment | 471.72 | 44 | <0.001 | 437.88 | 43 | <0.001 |
| WHR | Smoking initiation | 719.07 | 173 | <0.001 | 702.72 | 172 | <0.001 |
|  | Alcohol drinks per week | 563.69 | 173 | <0.001 | 560.08 | 172 | <0.001 |
|  | Risk tolerance | 496.94 | 198 | <0.001 | 486.63 | 197 | <0.001 |
|  | Educational attainment | 1429.33 | 195 | <0.001 | 1335.57 | 194 | <0.001 |
| T2D | Smoking initiation | 852.86 | 254 | <0.001 | 845.63 | 253 | <0.001 |
|  | Alcohol drinks per week | 852.40 | 256 | <0.001 | 850.76 | 255 | <0.001 |
|  | Risk tolerance | 556.93 | 273 | <0.001 | 551.33 | 272 | <0.001 |
|  | Educational attainment | 1893.23 | 271 | <0.001 | 1804.42 | 270 | <0.001 |
| HbA_1c_ | Smoking initiation | 93.86 | 36 | <0.001 | 91.43 | 35 | <0.001 |
|  | Alcohol drinks per week | 83.35 | 36 | <0.001 | 72.02 | 35 | <0.001 |
|  | Risk tolerance | 48.21 | 39 | 0.15 | 48.20 | 38 | 0.12 |
|  | Educational attainment | 249.64 | 39 | <0.001 | 249.48 | 38 | <0.001 |
| DBP | Smoking initiation | 207.72 | 63 | <0.001 | 206.47 | 62 | <0.001 |
|  | Alcohol drinks per week | 213.03 | 63 | <0.001 | 212.61 | 62 | <0.001 |
|  | Risk tolerance | 120.47 | 74 | <0.001 | 114.97 | 73 | <0.001 |
|  | Educational attainment | 308.47 | 73 | <0.001 | 293.92 | 72 | <0.001 |

Abbreviations: Q, Q-statistic; df, degrees of freedom; P, p-value; BMI, body mass index; WC, waist circumference; WHR, waist-hip ratio; T2D, type 2 diabetes mellitus; HbA_1c_, glycated haemoglobin; DBP, diastolic blood pressure.

**Table S11.** Assessing directional pleiotropy in Mendelian randomization results evaluating instrument-risk factor effects

| **Exposure** | **Outcome** | **MR Egger intercept estimate** | **SE** | **P** |
| --- | --- | --- | --- | --- |
| BMI | Smoking initiation | 0.003 | 0.001 | 0.01 |
|  | Alcohol drinks per week | 0.002 | 0.001 | 0.01 |
|  | Risk tolerance | 0.001 | 0.001 | 0.02 |
|  | Educational attainment | -0.003 | 0.001 | <0.001 |
| WC | Smoking initiation | 0.010 | 0.004 | 0.01 |
|  | Alcohol drinks per week | 0.003 | 0.001 | 0.07 |
|  | Risk tolerance | 0.002 | 0.002 | 0.19 |
|  | Educational attainment | -0.004 | 0.002 | 0.08 |
| WHR | Smoking initiation | 0.003 | 0.002 | 0.05 |
|  | Alcohol drinks per week | 0.001 | 0.001 | 0.29 |
|  | Risk tolerance | 0.001 | 0.001 | 0.04 |
|  | Educational attainment | -0.004 | 0.001 | <0.001 |
| T2D | Smoking initiation | 0.001 | 0.001 | 0.14 |
|  | Alcohol drinks per week | -0.001 | 0.001 | 0.48 |
|  | Risk tolerance | 0.001 | 0.001 | 0.10 |
|  | Educational attainment | -0.002 | 0.001 | <0.001 |
| HbA_1c_ | Smoking initiation | -0.002 | 0.002 | 0.34 |
|  | Alcohol drinks per week | -0.002 | 0.001 | 0.02 |
|  | Risk tolerance | 0.001 | 0.001 | 0.92 |
|  | Educational attainment | -0.001 | 0.001 | 0.88 |
| DBP | Smoking initiation | -0.002 | 0.003 | 0.54 |
|  | Alcohol drinks per week | 0.001 | 0.002 | 0.73 |
|  | Risk tolerance | -0.002 | 0.001 | 0.07 |
|  | Educational attainment | -0.003 | 0.002 | 0.06 |

Abbreviations: SE, standard error; P, p-value; BMI, body mass index; WC, waist circumference; WHR, waist-hip ratio; T2D, type 2 diabetes mellitus; HbA_1c_, glycated haemoglobin; DBP, diastolic blood pressure.

**Table S12.** Outlier corrected Mendelian randomization results evaluating instrument-risk factor effects

| **Exposure** | **Outcome** | **N Outlier SNPs** | **Beta** | **SE** | **P** |
| --- | --- | --- | --- | --- | --- |
| BMI | Smoking initiation | 64 | 0.23 | 0.02 | <0.001 |
|  | Alcohol drinks per week | 80 | -0.01 | 0.01 | 0.41 |
|  | Risk tolerance | 74 | 0.04 | 0.01 | <0.001 |
|  | Educational attainment | 99 | -0.16 | 0.01 | <0.001 |
| WC | Smoking initiation | 24 | 0.17 | 0.04 | <0.001 |
|  | Alcohol drinks per week | 14 | -0.09 | 0.01 | <0.001 |
|  | Risk tolerance | 11 | 0.01 | 0.02 | 0.50 |
|  | Educational attainment | 19 | -0.06 | 0.02 | <0.001 |
| WHR | Smoking initiation | 52 | 0.15 | 0.02 | <0.001 |
|  | Alcohol drinks per week | 39 | -0.02 | 0.01 | 0.12 |
|  | Risk tolerance | 36 | 0.02 | 0.01 | 0.02 |
|  | Educational attainment | 75 | -0.11 | 0.01 | <0.001 |
| T2D | Smoking initiation | 66 | 0.01 | 0.01 | 0.01 |
|  | Alcohol drinks per week | 49 | -0.02 | 0.01 | <0.001 |
|  | Risk tolerance | 42 | 0.01 | 0.01 | 0.30 |
|  | Educational attainment | 101 | -0.01 | 0.01 | <0.001 |
| HbA_1c_ | Smoking initiation | 6 | 0.02 | 0.04 | 0.64 |
|  | Alcohol drinks per week | 8 | 0.01 | 0.03 | 0.62 |
|  | Risk tolerance | 1 | 0.01 | 0.02 | 0.96 |
|  | Educational attainment | 11 | -0.07 | 0.02 | <0.001 |
| DBP | Smoking initiation | 13 | 0.01 | 0.01 | 0.91 |
|  | Alcohol drinks per week | 17 | 0.01 | 0.01 | 0.30 |
|  | Risk tolerance | 9 | 0.01 | 0.01 | 0.39 |
|  | Educational attainment | 25 | 0.01 | 0.01 | 0.66 |

Abbreviations: IVW, inverse variance weighted; OR, odds ratio; CI, confidence intervals; P, p-value; BMI, body mass index; WC, waist circumference; WHR, waist-hip ratio; T2D, type 2 diabetes mellitus;, glycated haemoglobin; DBP, diastolic blood pressure. OR are expressed per 1 standard deviation (SD) increase in genetically predicted BMI (4.81 kg/m2), WC (0.09 unit), WHR (0.10 unit), T2D (1-log unit higher odds of T2D), HbA1c (1-log-unit % higher glycated haemoglobin), and DBP (1 unit mmHg increase). Outcome beta estimates reflect the standard deviation of the phenotype.

**Figures S1A-C.** Power calculations for oral and oropharyngeal analyses in GAME-ON.

A. Combined oral and oropharyngeal cancer cases

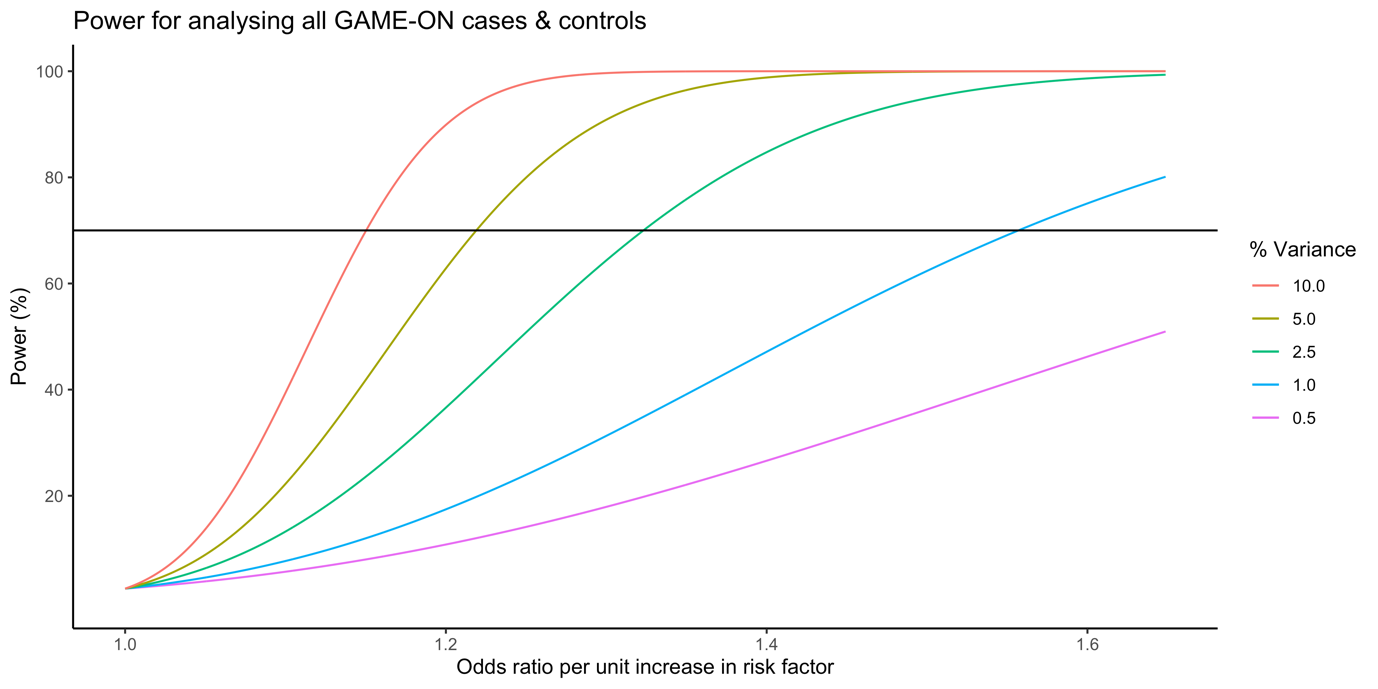

B. Oral cancer cases only

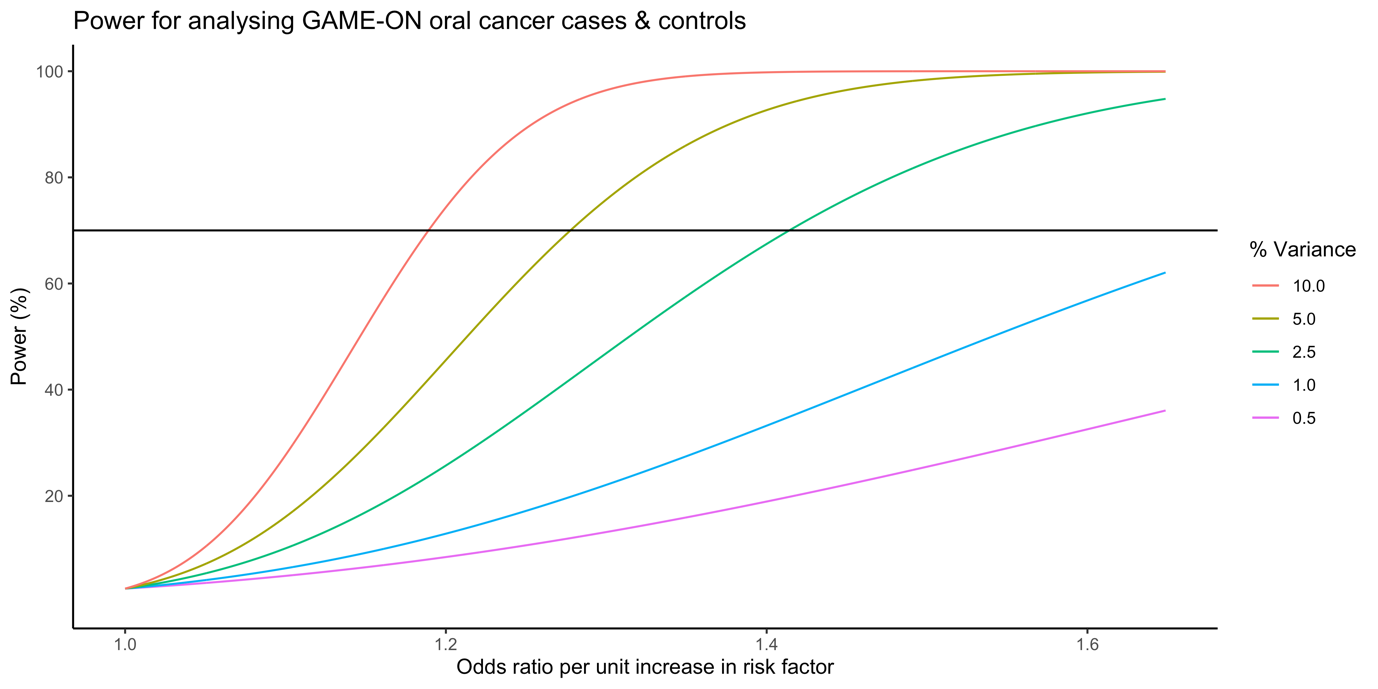

C. Oropharyngeal cancer cases only

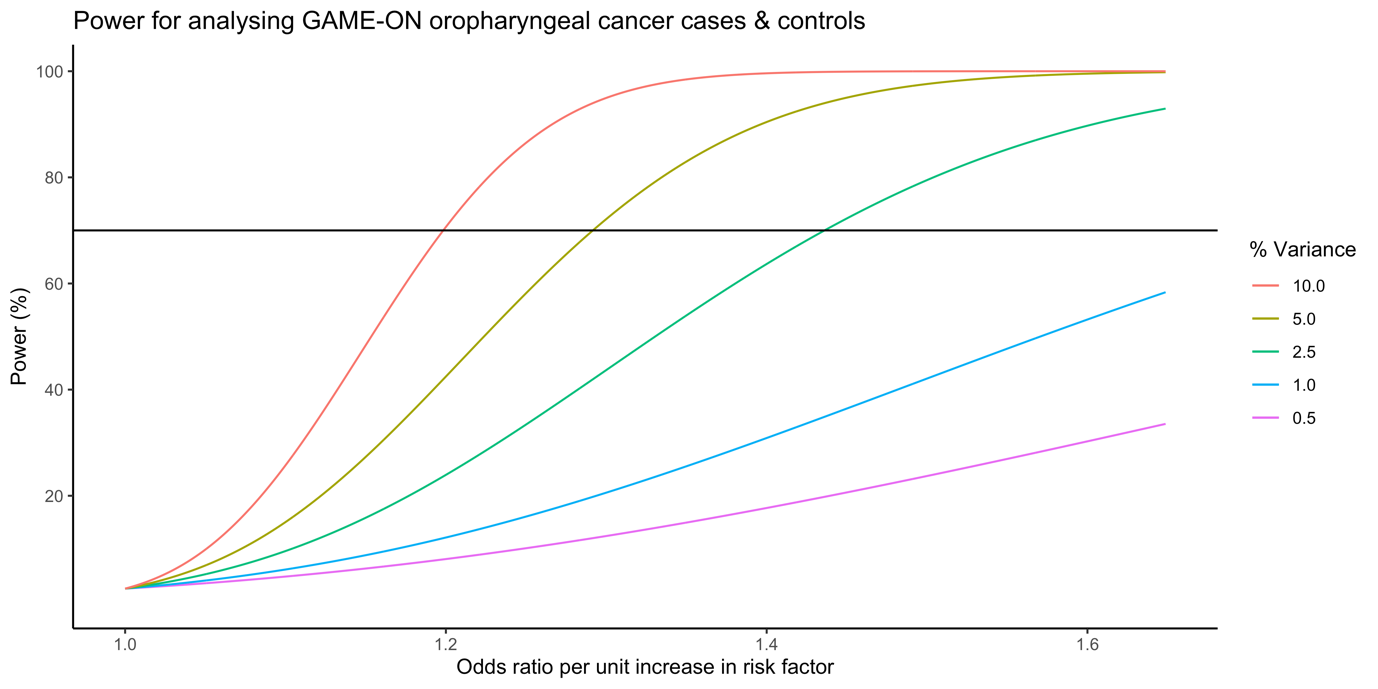

Alpha α set at 0.05.

**Figure S2. Forest plots showing Mendelian randomization results for genetically-proxied body mass index (BMI) with risk of combined oral and oropharyngeal cancer in GAME-ON.** Effect estimates on oral and oropharyngeal cancer are reported on the log odds scale.

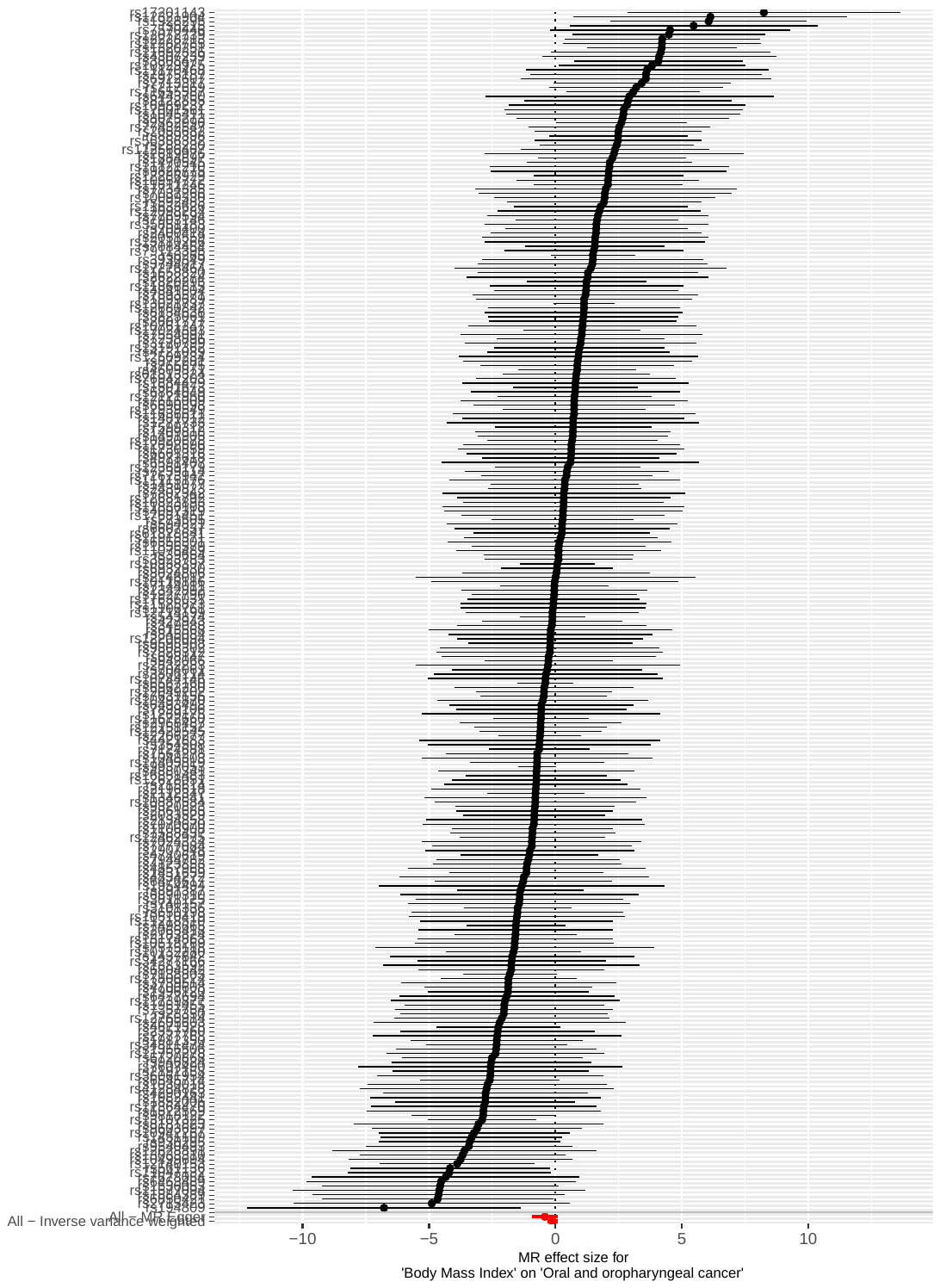

**Figure S3. Forest plots showing Mendelian randomization results for genetically-proxied waist hip ratio (WHR) with risk of combined oral and oropharyngeal cancer in GAME-ON.** Effect estimates on oral and oropharyngeal cancer are reported on the log odds scale.

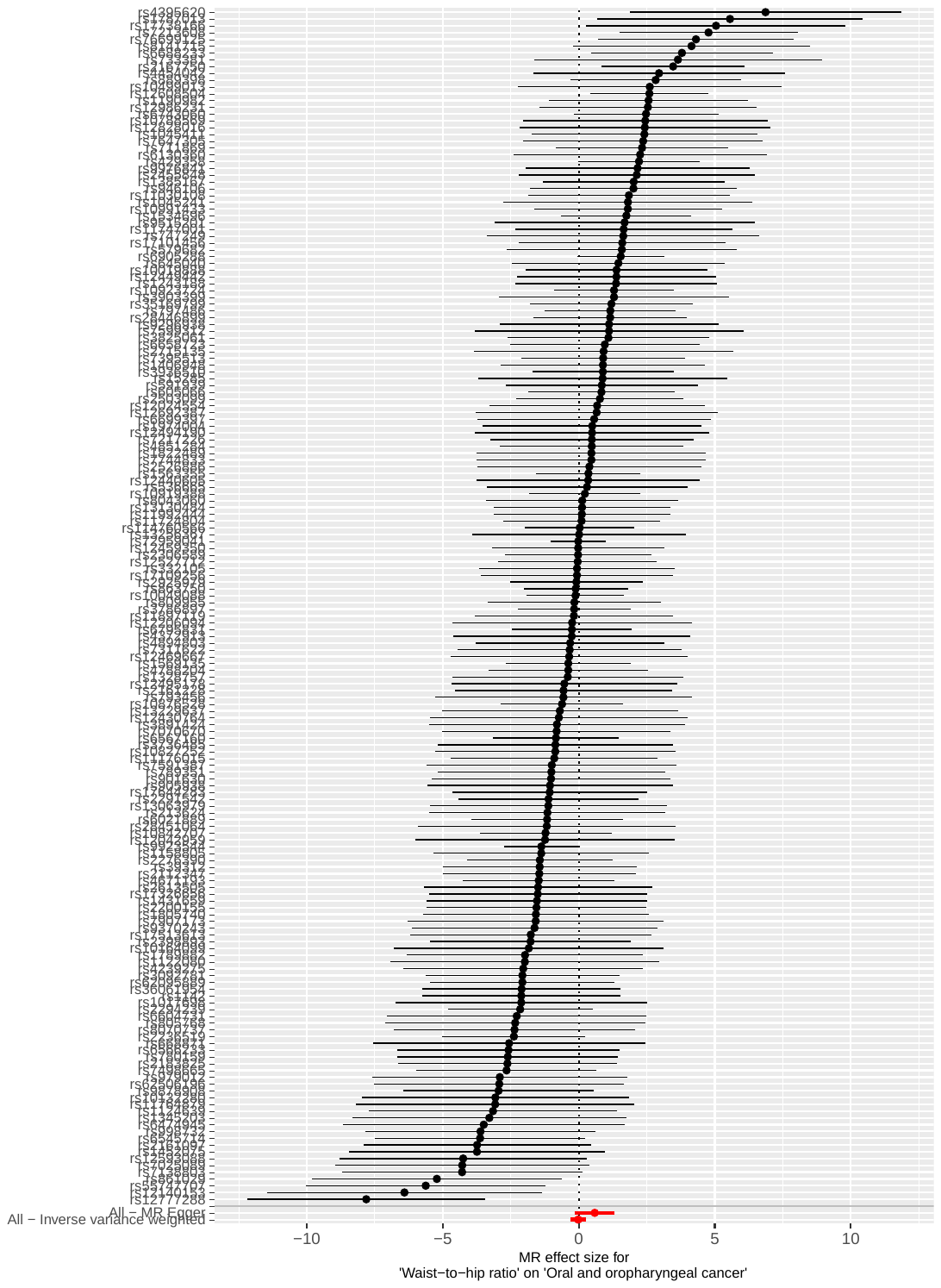

**Figure S4. Forest plots showing Mendelian randomization results for genetically-proxied waist circumference (WC) with risk of combined oral and oropharyngeal cancer in GAME-ON.** Effect estimates on oral and oropharyngeal cancer are reported on the log odds scale.

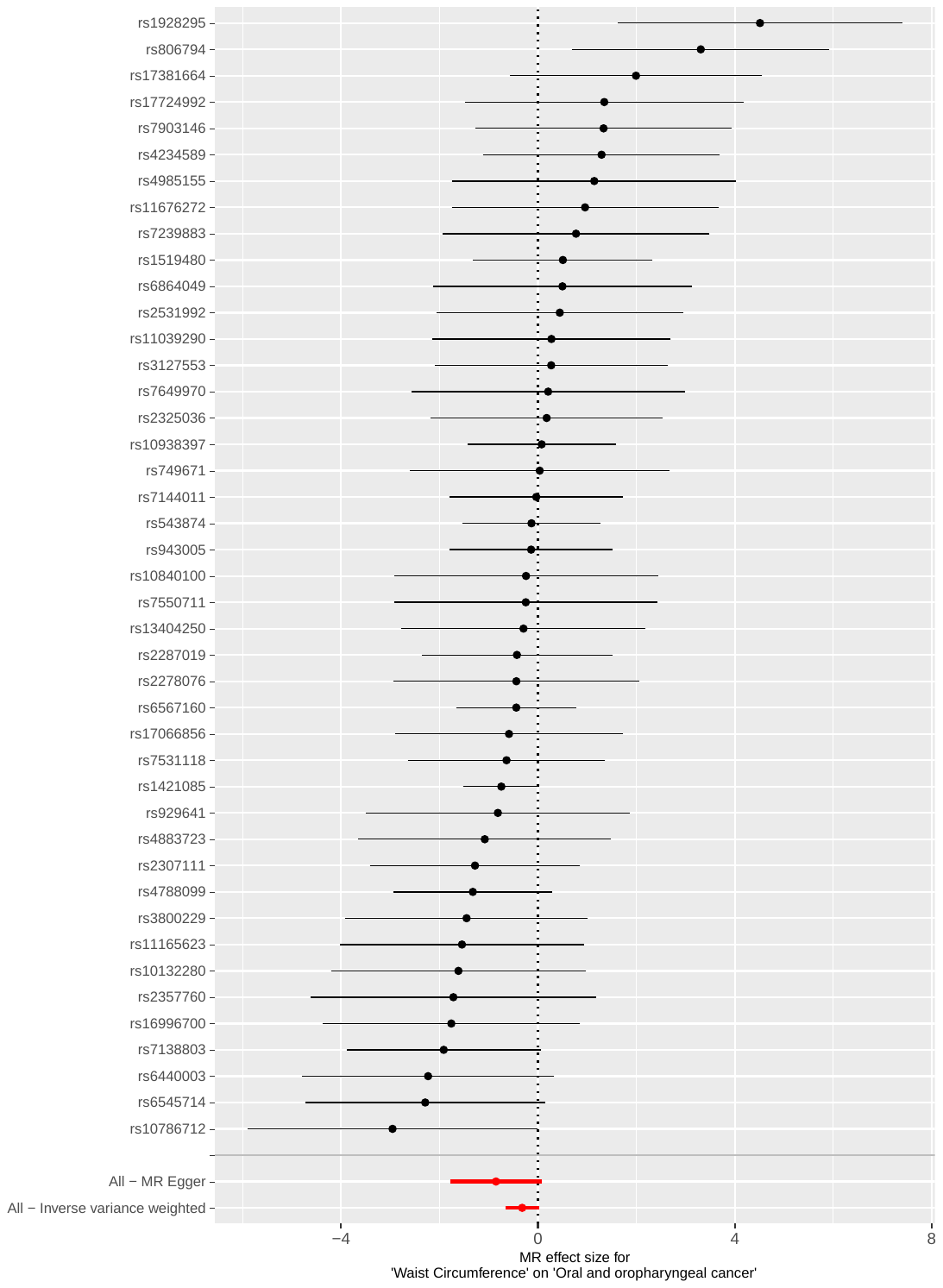

**Figure S5. Forest plots showing Mendelian randomization results for genetically-proxied type 2 diabetes mellitus (T2D) with risk of combined oral and oropharyngeal cancer in GAME-ON.** Effect estimates on oral and oropharyngeal cancer are reported on the log odds scale.

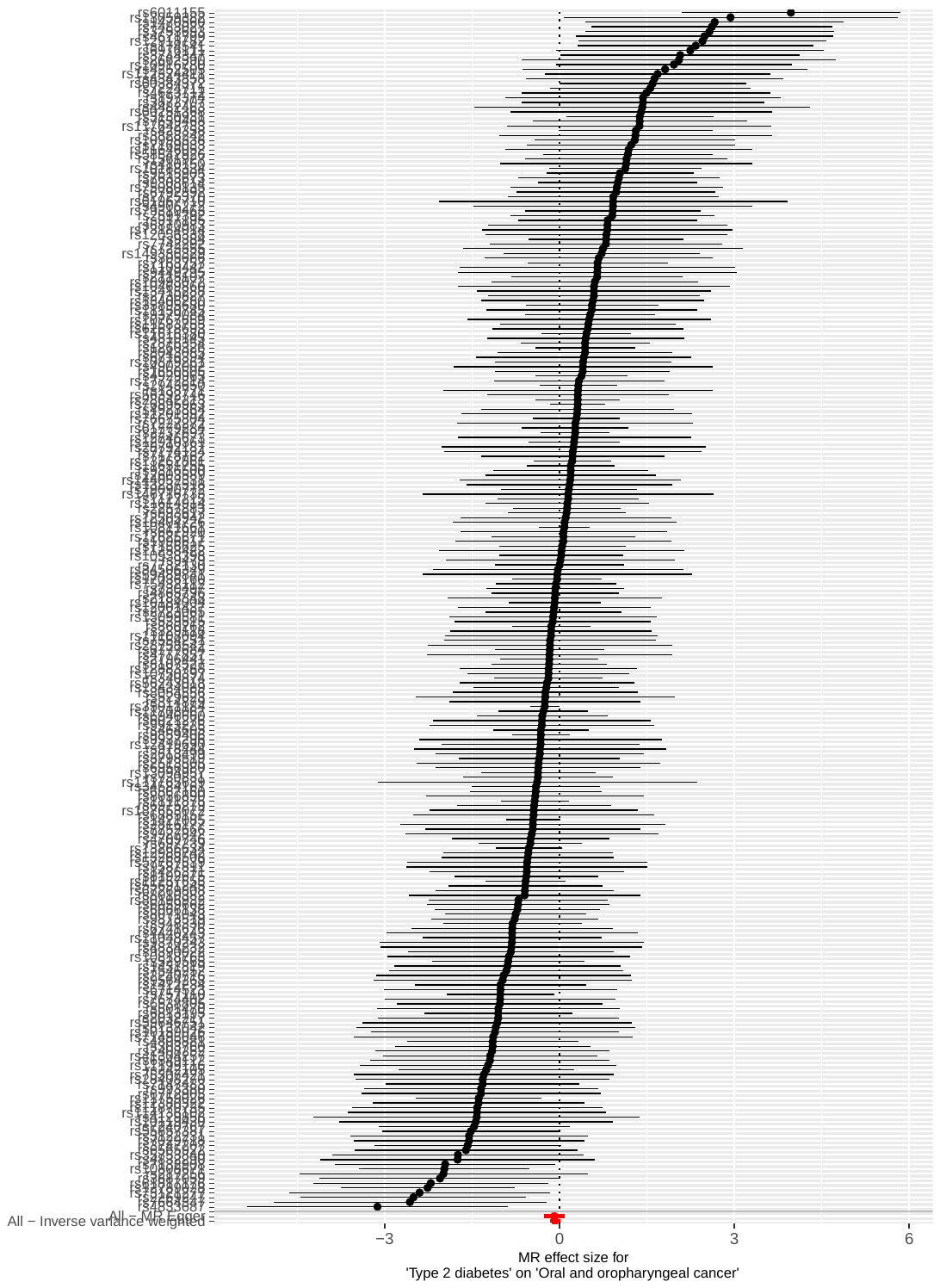

**Figure S6. Forest plots showing Mendelian randomization results for genetically-proxied glycated haemoglobin (HbA_1c_) with risk of combined oral and oropharyngeal cancer in GAME-ON.** Effect estimates on oral and oropharyngeal cancer are reported on the log odds scale.

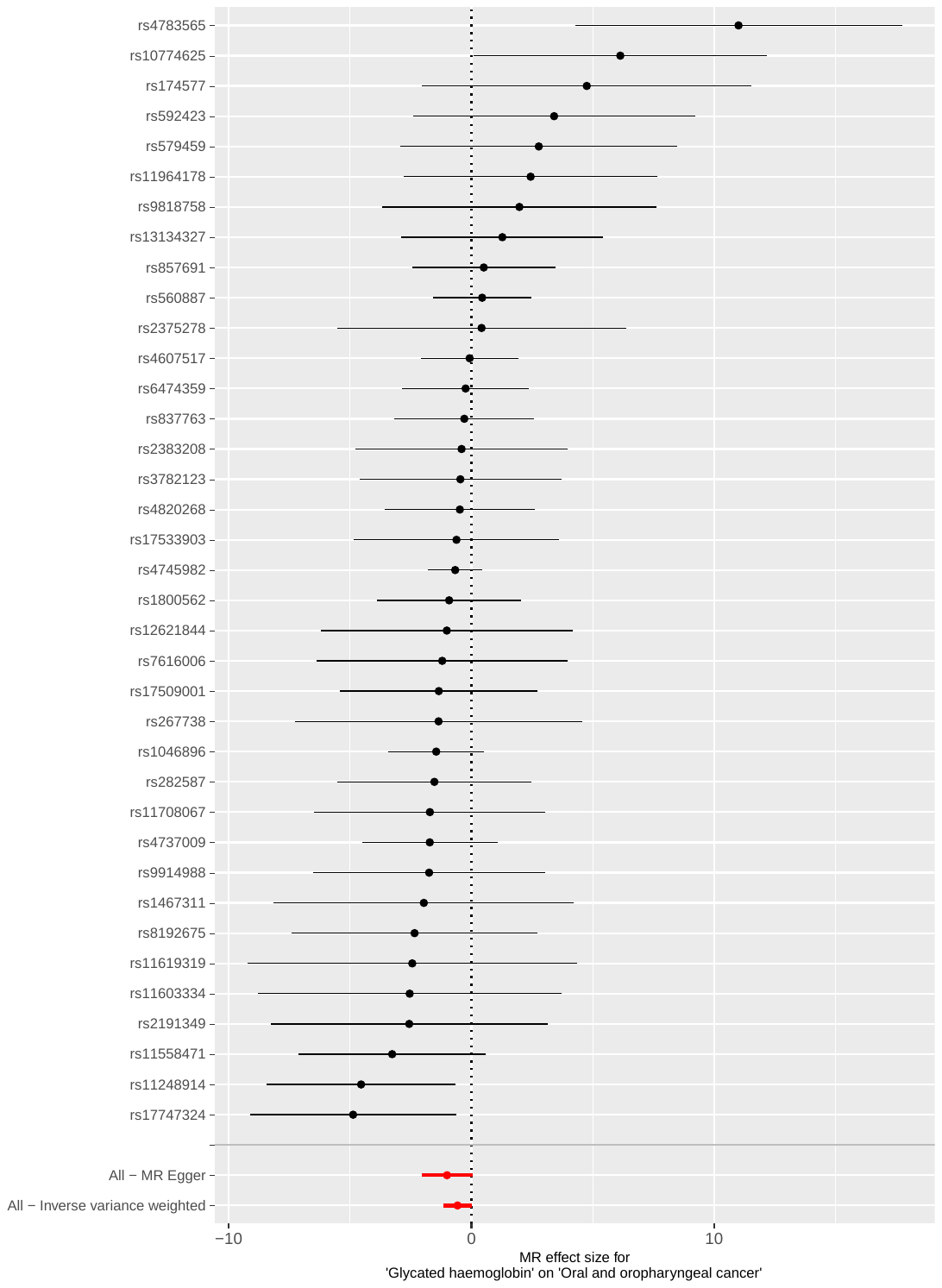

**Figure S7. Forest plots showing Mendelian randomization results for genetically-proxied fasting glucose (FG) with risk of combined oral and oropharyngeal cancer in GAME-ON.** Effect estimates on oral and oropharyngeal cancer are reported on the log odds scale.

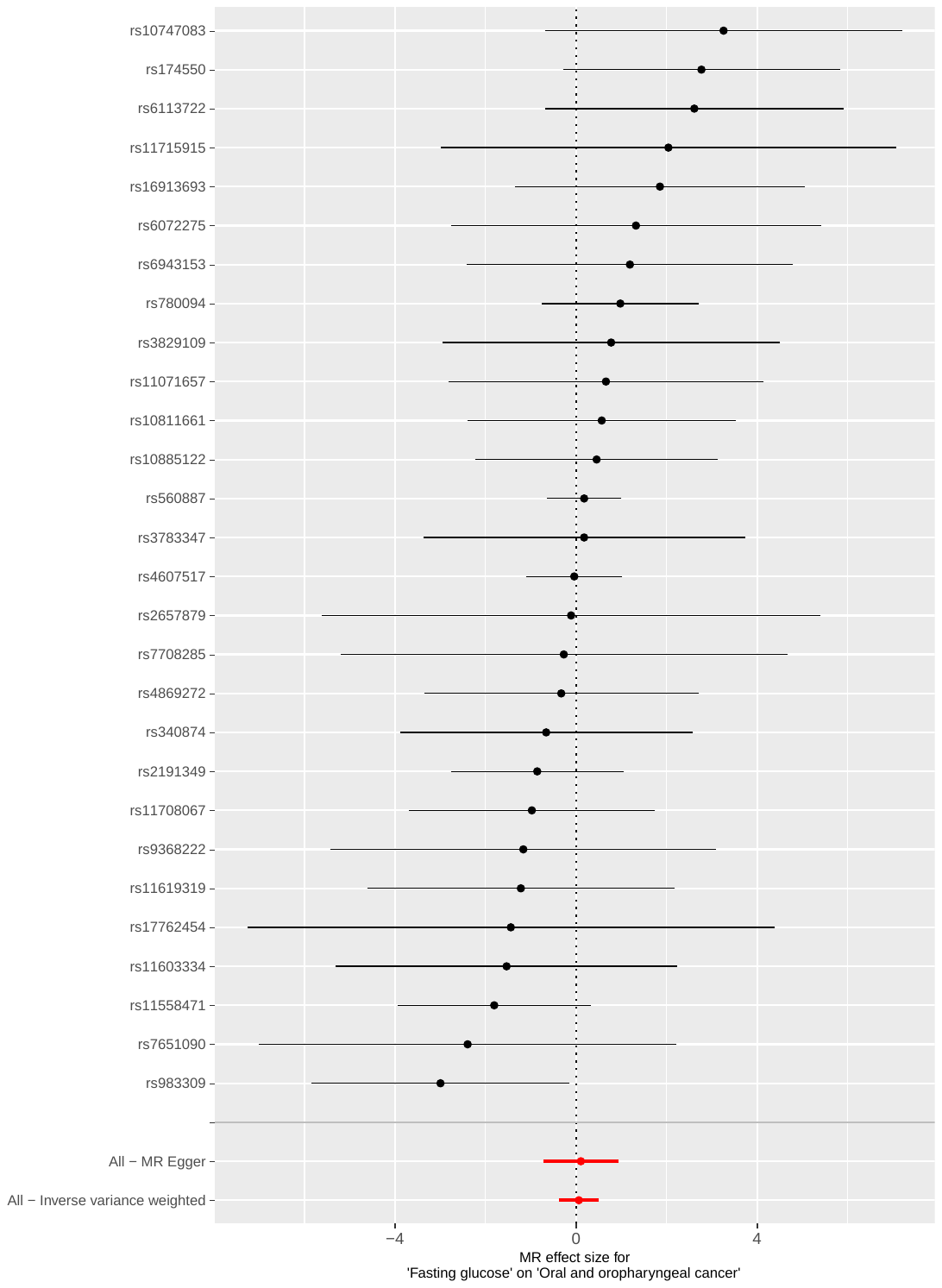

**Figure S8. Forest plots showing Mendelian randomization results for genetically-proxied fasting insulin (FI) with risk of combined oral and oropharyngeal cancer in GAME-ON.** Effect estimates on oral and oropharyngeal cancer are reported on the log odds scale.

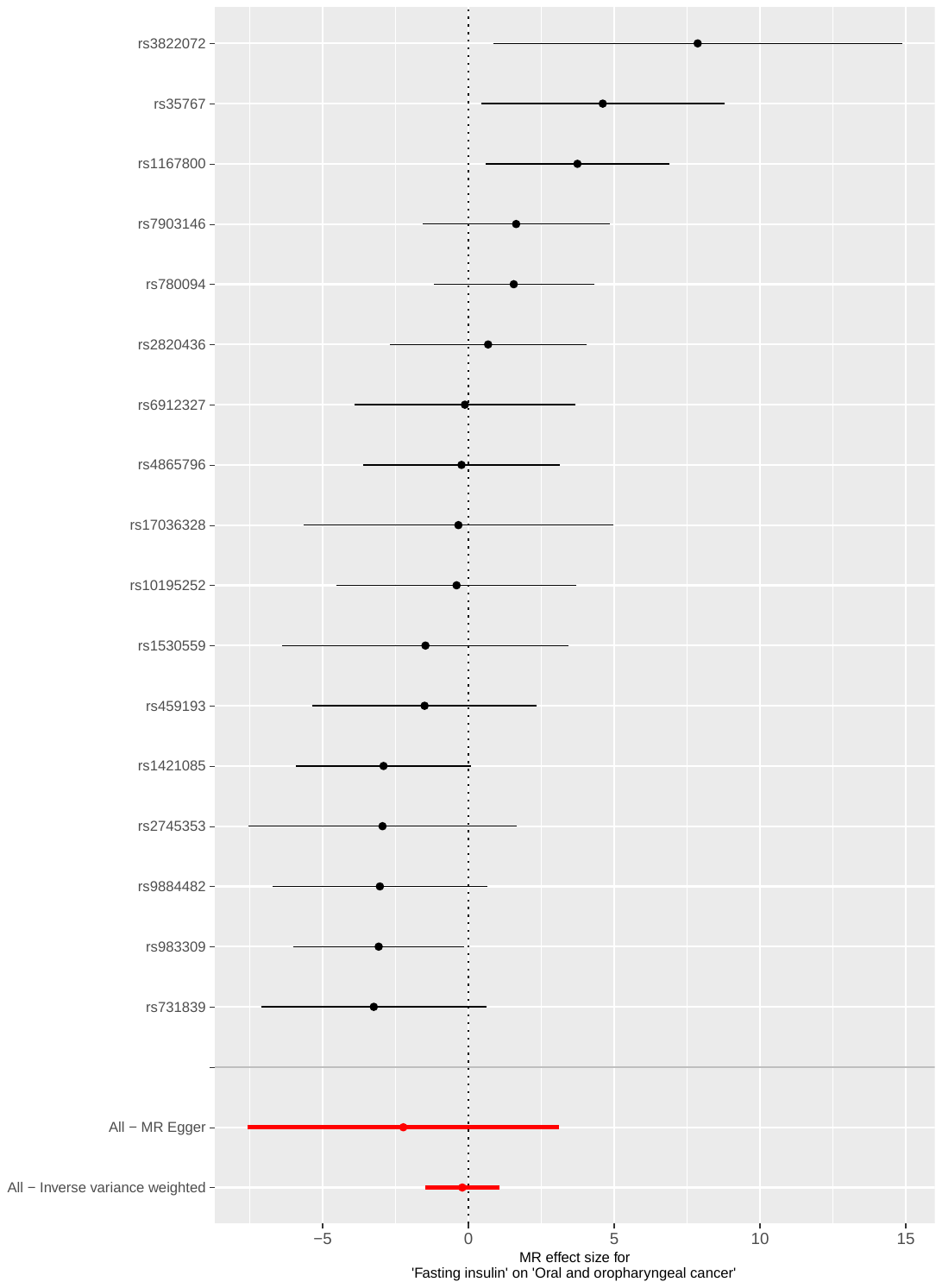

**Figure S9. Forest plots showing Mendelian randomization results for genetically-proxied systolic blood pressure (SBP) with risk of combined oral and oropharyngeal cancer in GAME-ON.** Effect estimates on oral and oropharyngeal cancer are reported on the log odds scale.

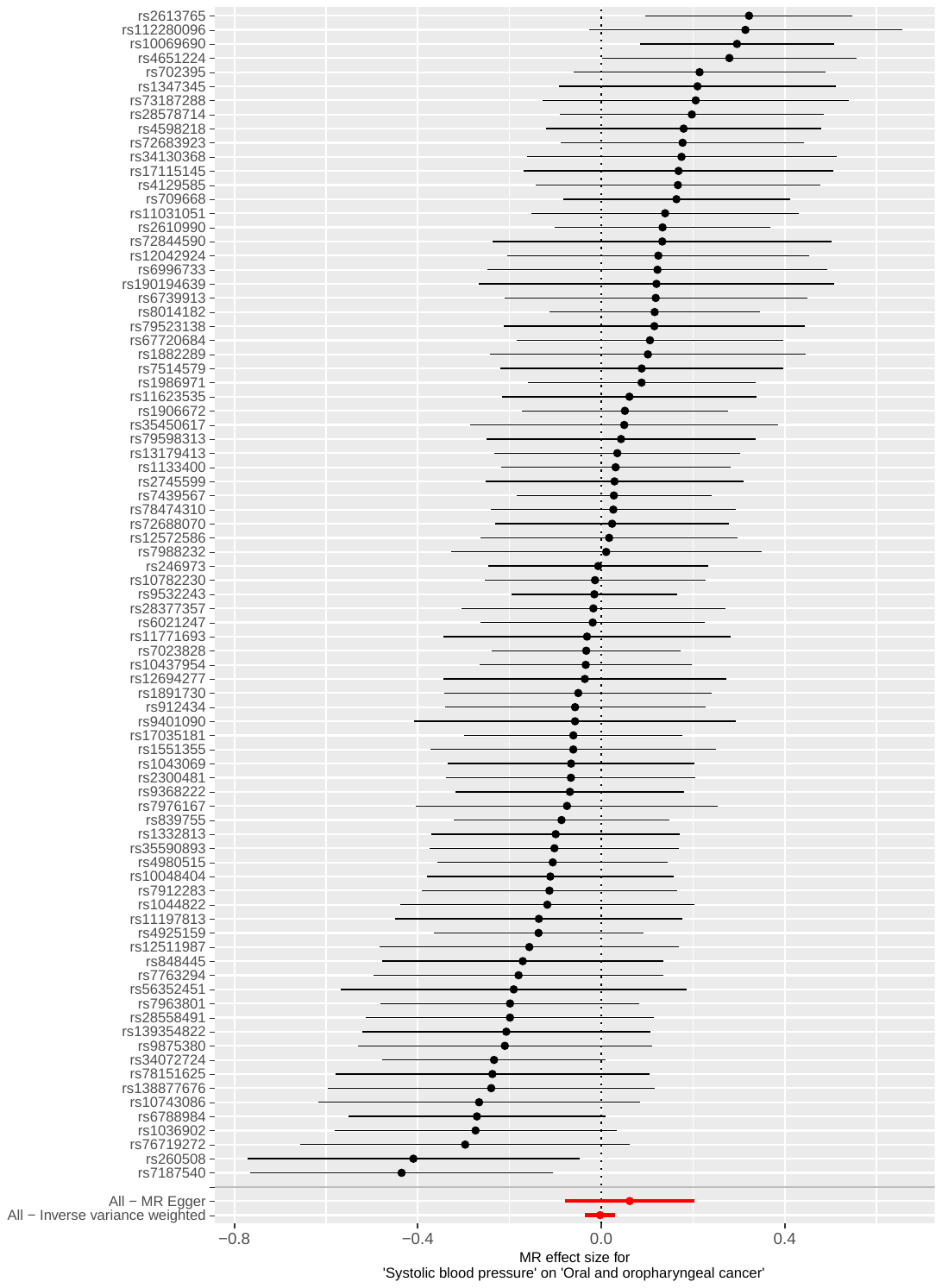

**Figure S10. Forest plots showing Mendelian randomization results for genetically-proxied diastolic blood pressure (DBP) with risk of combined oral and oropharyngeal cancer in GAME-ON.** Effect estimates on oral and oropharyngeal cancer are reported on the log odds scale.

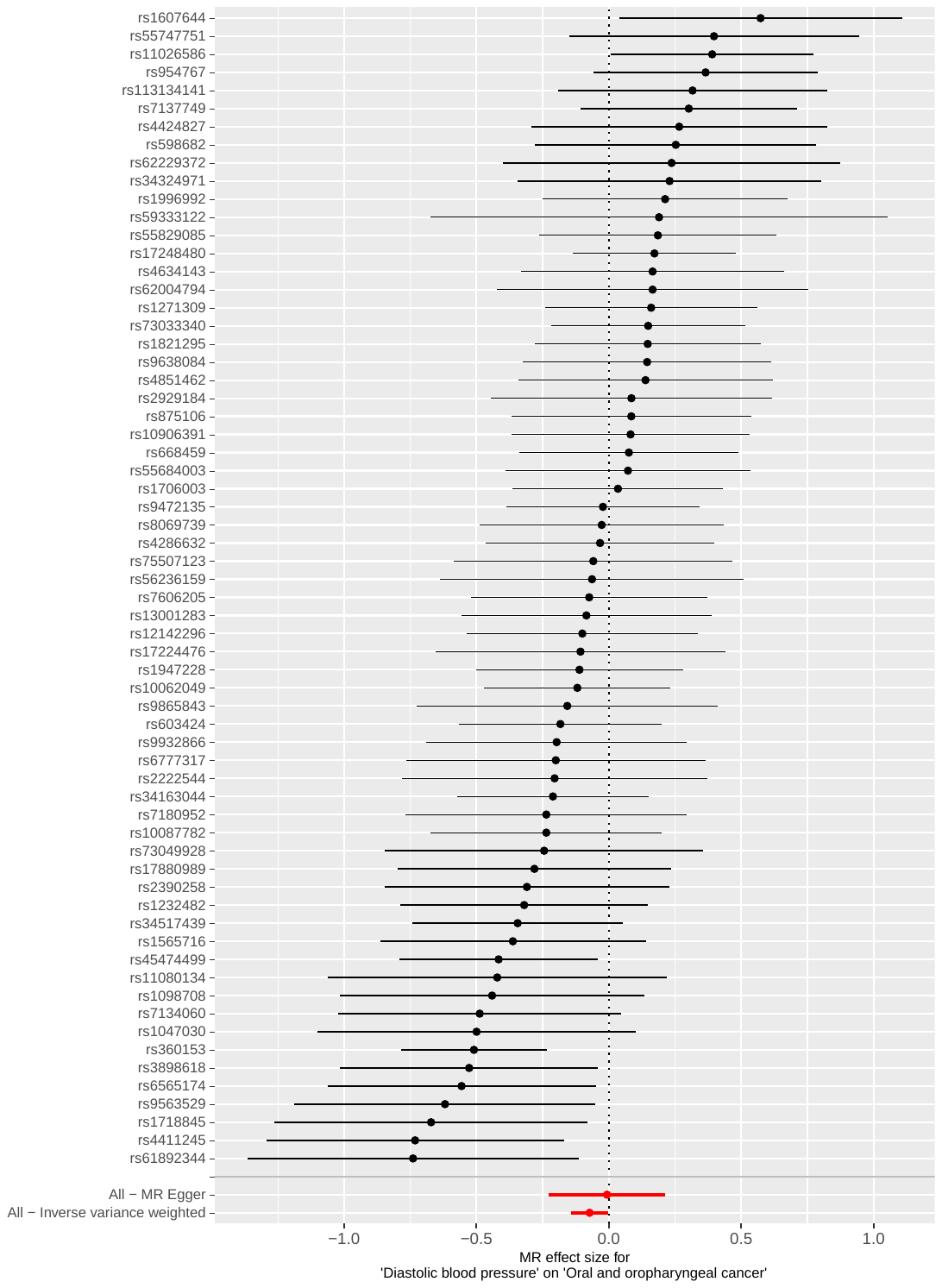

**Figure S11. Scatter plot for body mass index (BMI) with risk of combined oral and oropharyngeal cancer in GAME-ON.**

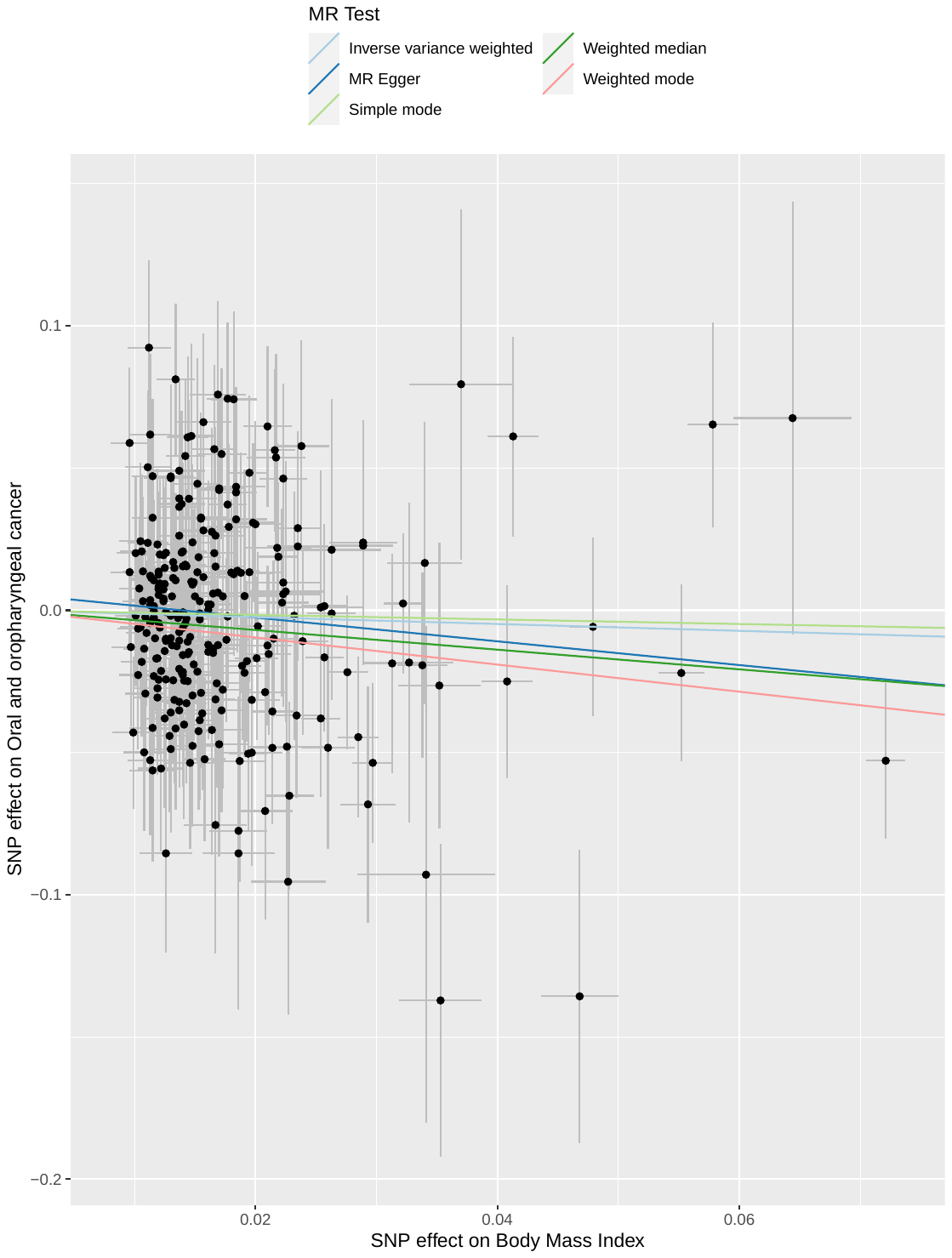

**Figure S12. Scatter plot for waist circumference (WC) with risk of combined oral and oropharyngeal cancer in GAME-ON.**

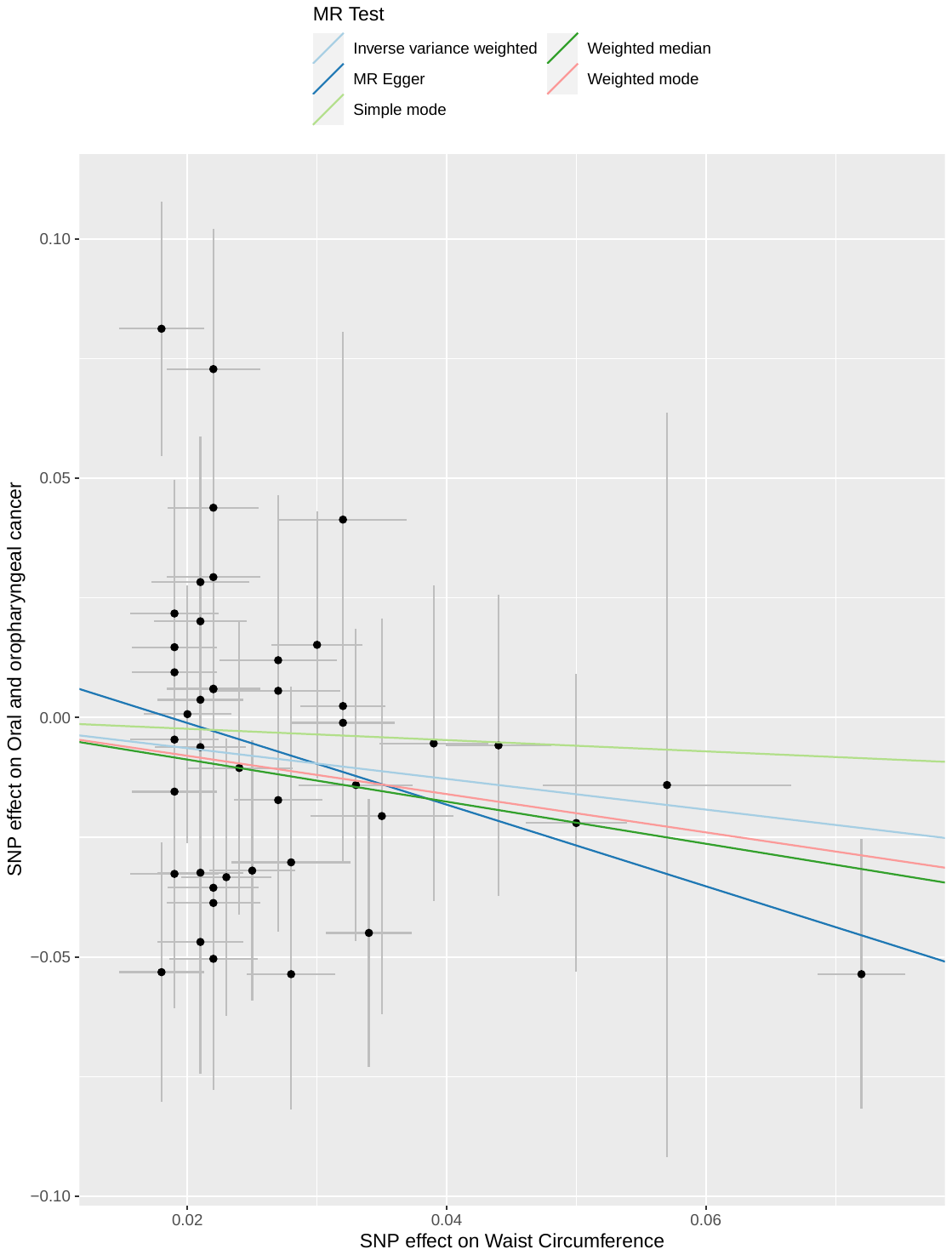

**Figure S13. Scatter plot for waist hip ratio (WHR) with risk of combined oral and oropharyngeal cancer in GAME-ON.**

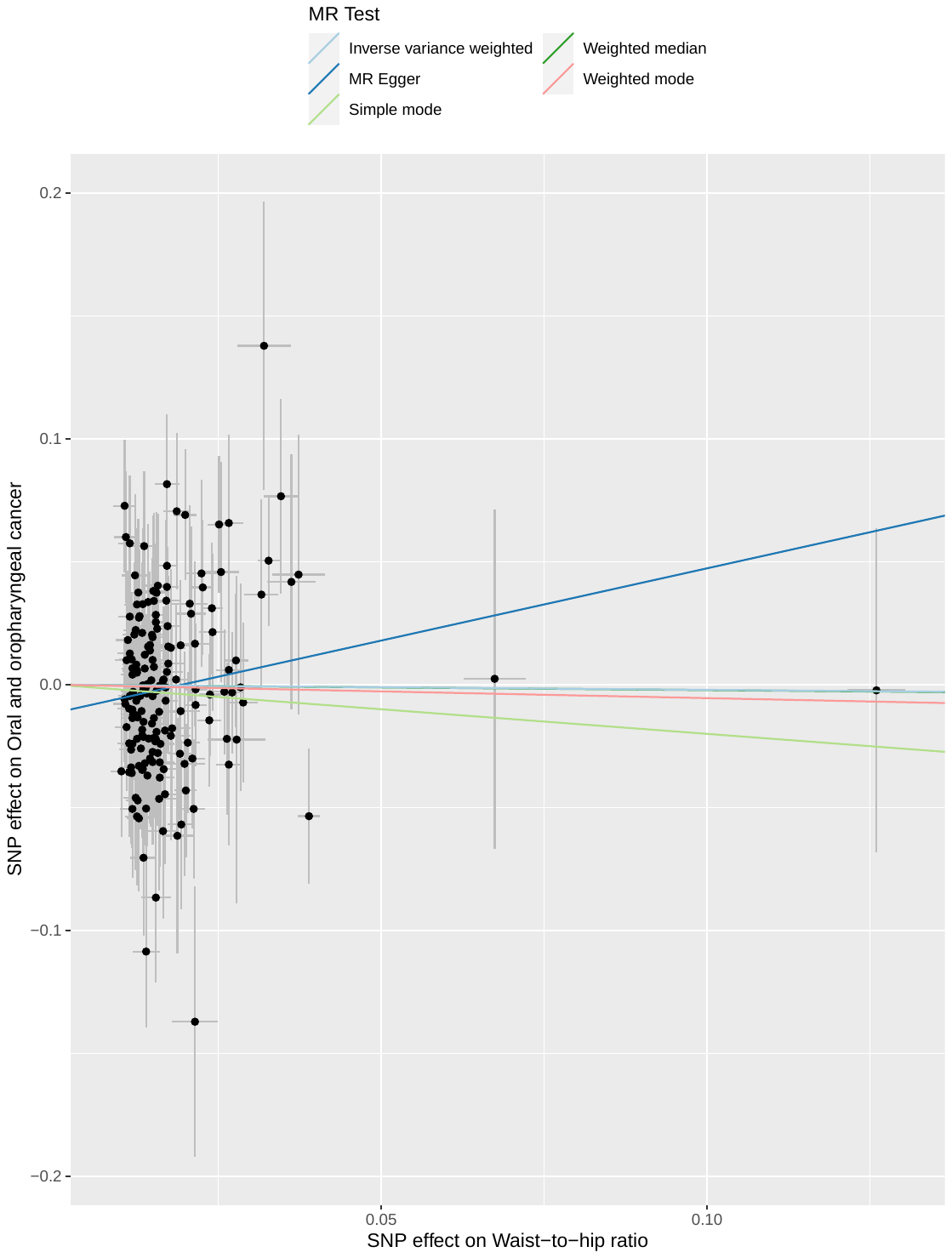

**Figure S14. Scatter plot for type 2 diabetes mellitus (T2D) with risk of combined oral and oropharyngeal cancer in GAME-ON.**

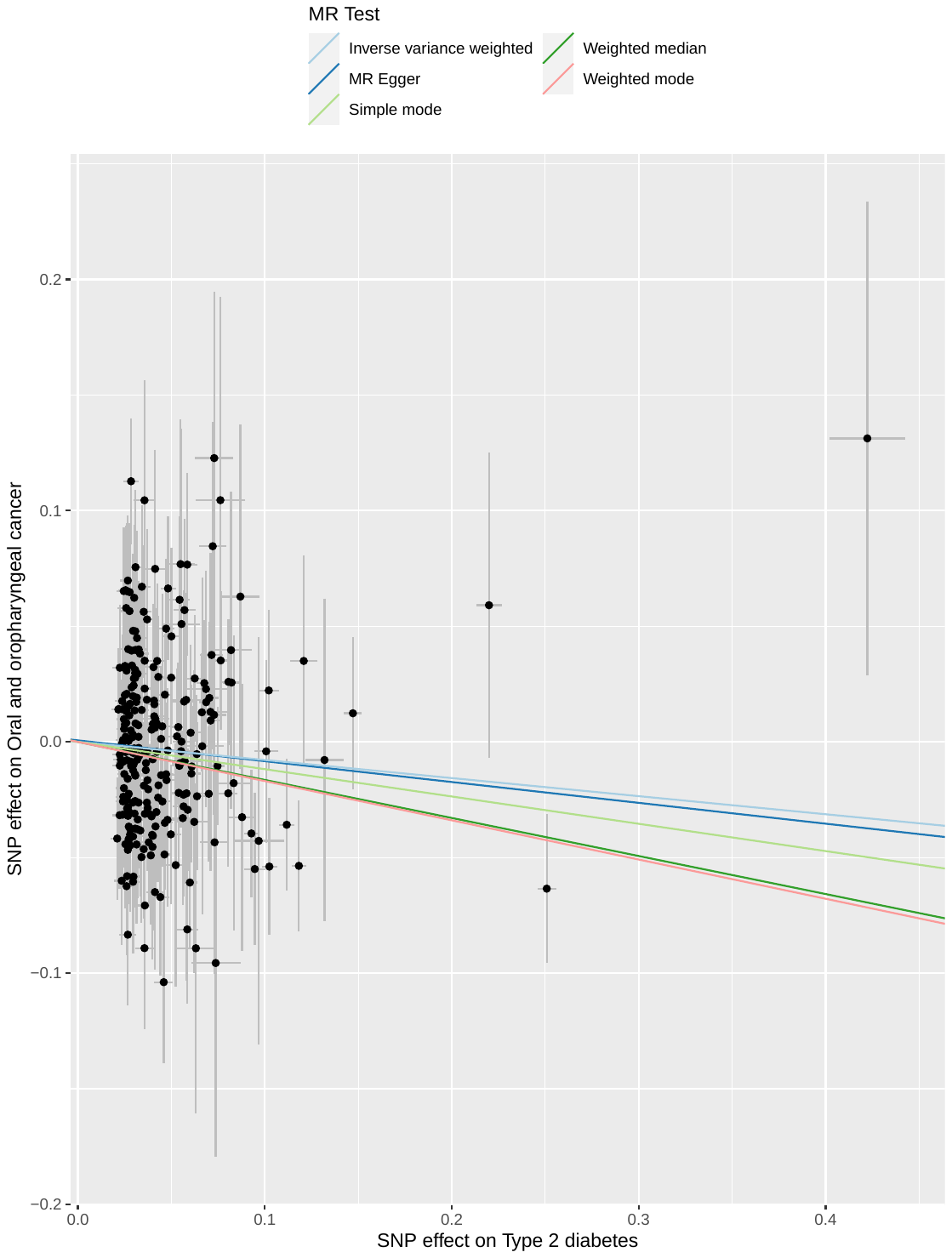

**Figure S15. Scatter plot for glycated haemoglobin (HBA_1c_) with risk of combined oral and oropharyngeal cancer in GAME-ON.**

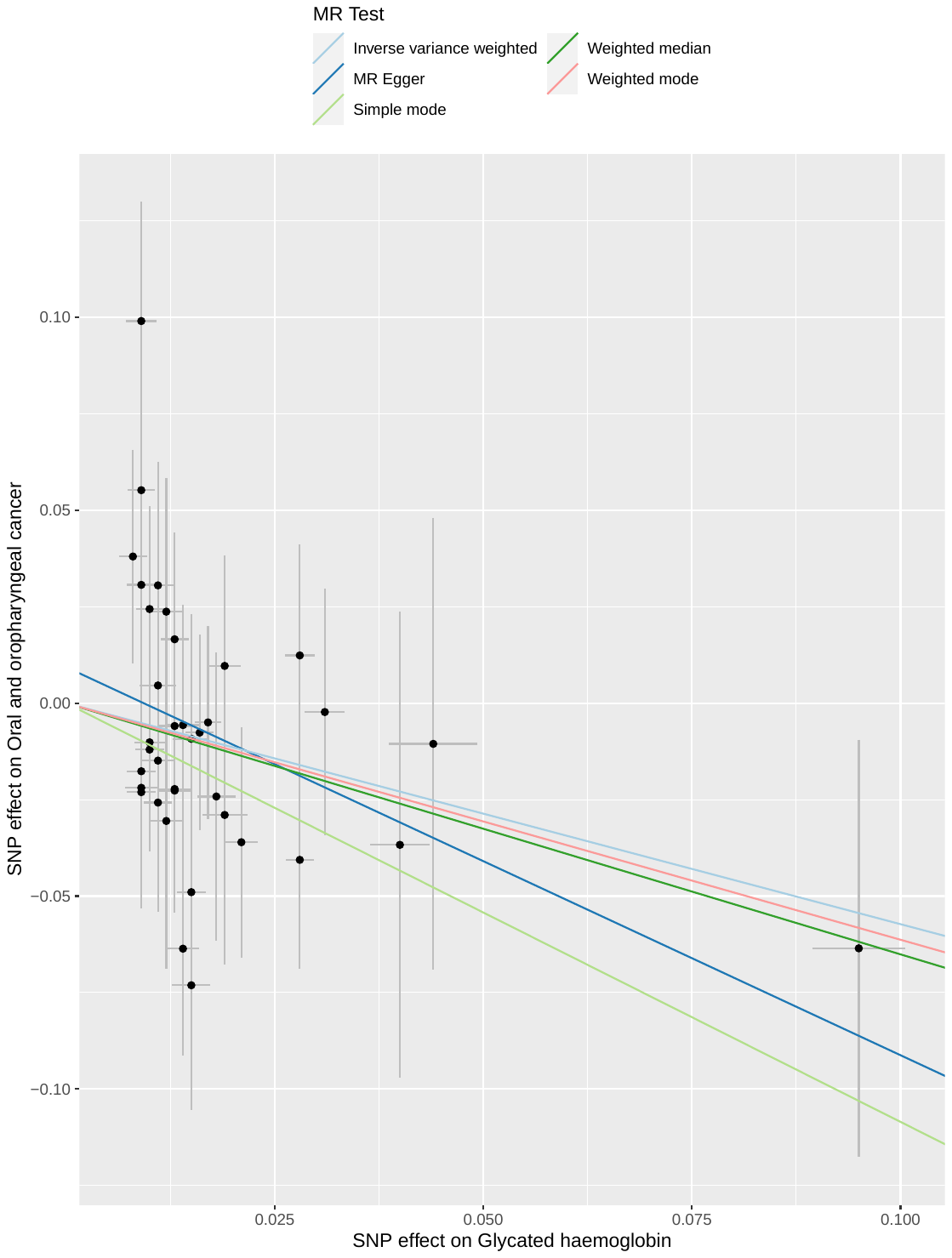

**Figure S16. Scatter plot for fasting glucose (FG) with risk of combined oral and oropharyngeal cancer in GAME-ON.**

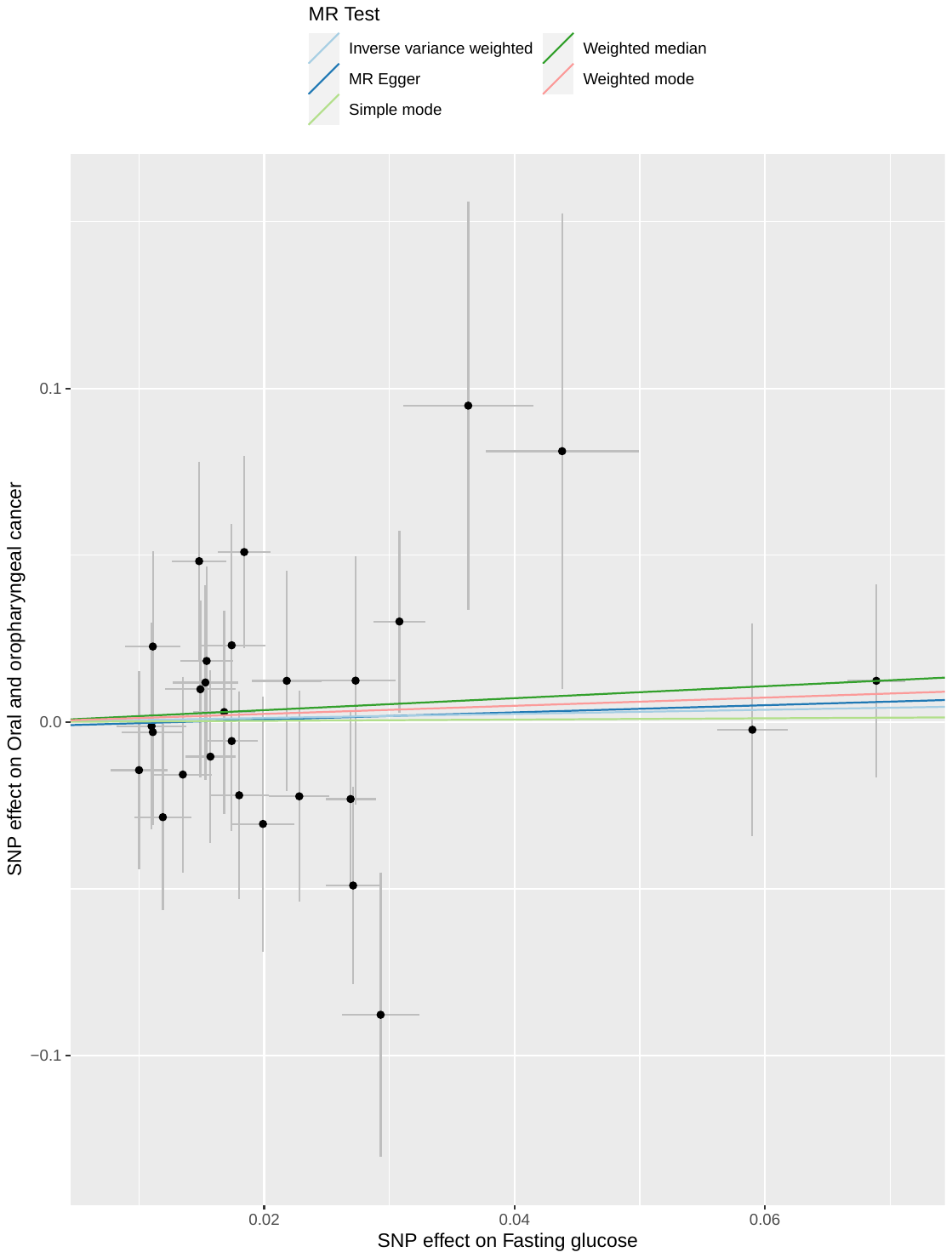

**Figure S17. Scatter plot for fasting insulin (FI) with risk of combined oral and oropharyngeal cancer in GAME-ON.**

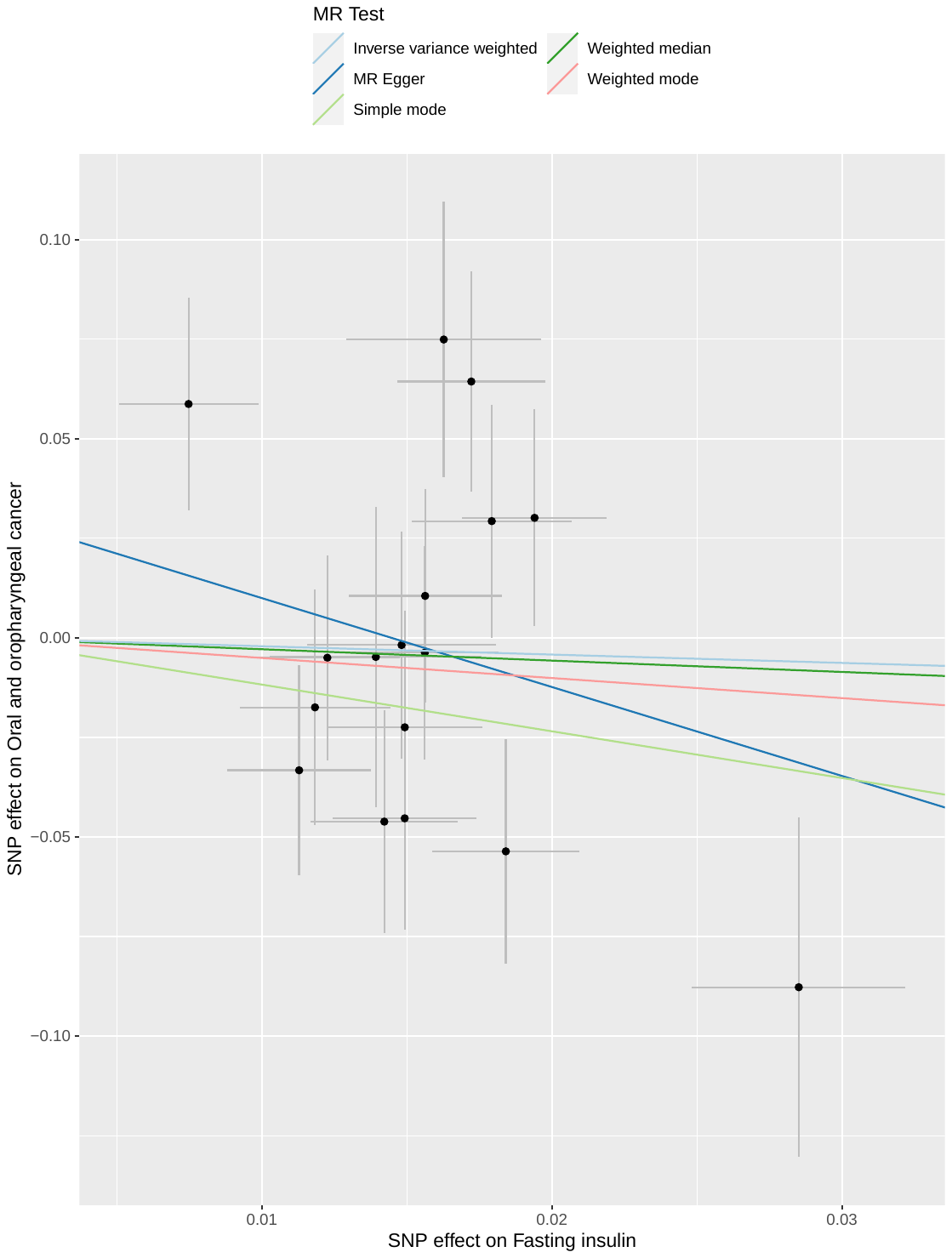

**Figure S18. Scatter plot for systolic blood pressure (SBP) with risk of combined oral and oropharyngeal cancer in GAME-ON.**

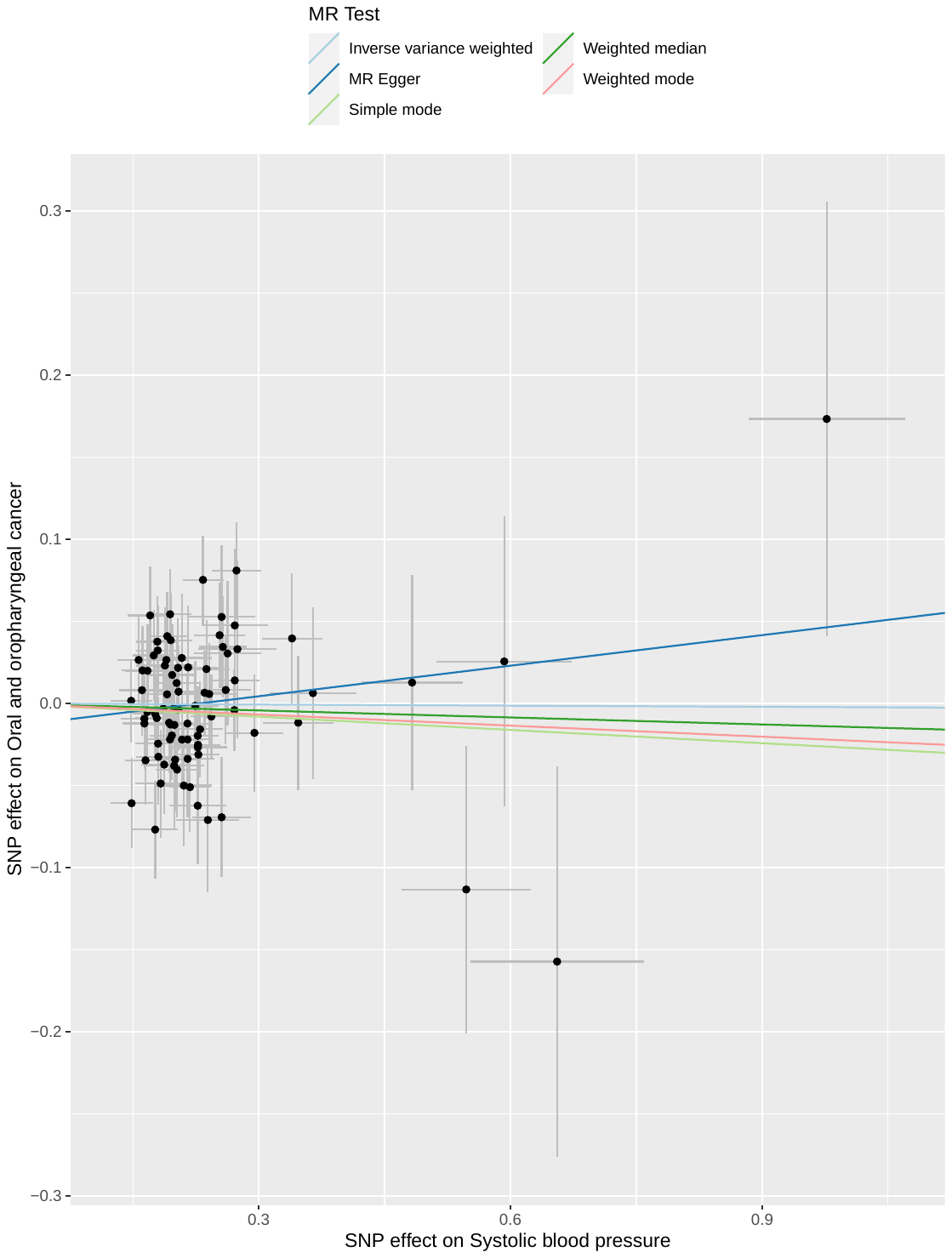

**Figure S19. Scatter plot for diastolic blood pressure (DBP) with risk of combined oral and oropharyngeal cancer in GAME-ON.**

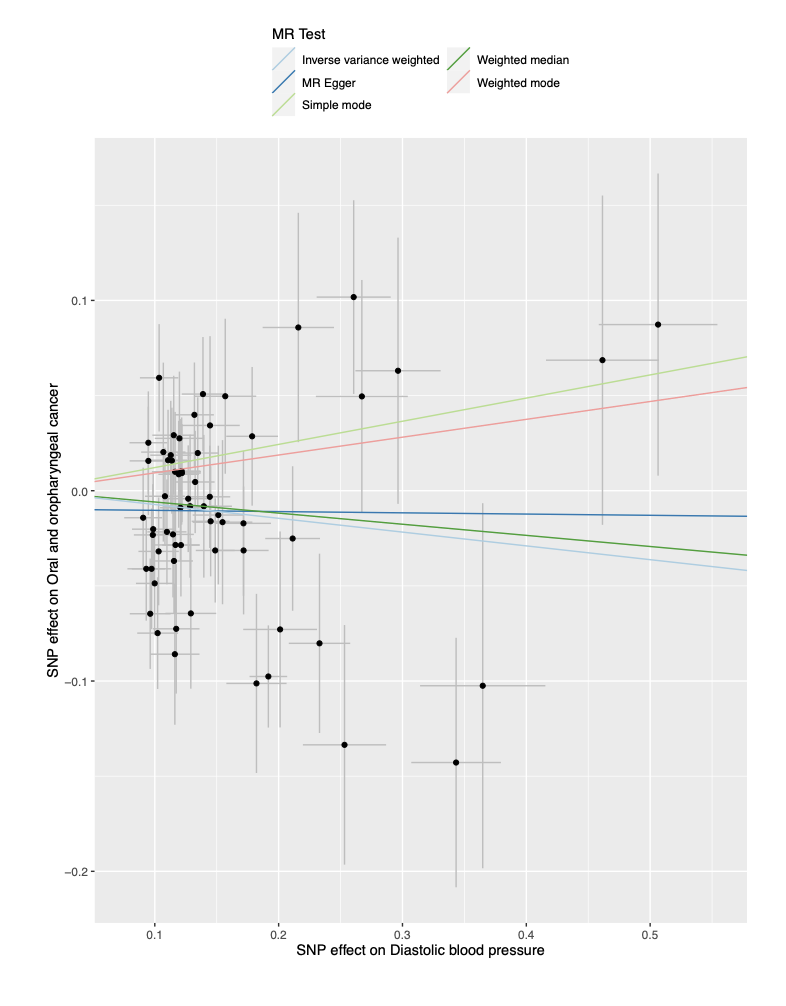
