## Supplementary material for "Evaluating the effect of metabolic traits on oral and oropharyngeal cancer risk using Mendelian randomization": STROBE MR checklist

**Supplementary File 2. STROBE-MR checklist of recommended items to address in reports of *Mendelian randomization studies***

|  | **Item Nº** | **Recommendation** | **Comments/page Nº** |
| --- | --- | --- | --- |
| **Title and abstract** |  |  |  |
|  | 1 | Indicate Mendelian randomization (MR) as the study’s design in the title and/or the abstract if that is a main purpose of the study. | ‘Mendelian randomization’ in title and abstract |
| **Introduction** |  |  |  |
| Background | 2 | Explain the scientific background and rationale for the reported study. Is a potential causal relationship between exposure and outcome plausible? Justify why MR is a helpful method to address the study question. | Pages 3–5 |
| Objectives | 3 | State specific objectives clearly, including pre-specified causal hypotheses (if any). State that MR is a method that, under specific assumptions, intends to estimate causal effects. | Page 5 |
| **Methods** |  |  |  |
| Study design and data sources | 4 | Present key elements of study design early in the article. Consider including a table listing sources of data for all phases of the study. For each data source contributing to the analysis, describe the following:  (*a*) Setting: Describe the study design and the underlying population, if possible. Describe the setting, locations, and relevant dates, including periods of recruitment, exposure, follow-up, and data collection, when available.  (*b*) Participants: Give the eligibility criteria, and the sources and methods of selection of participants. Report the sample size, and whether any power or sample size calculations were carried out prior to the main analysis.  c) Explain how the analysed sample size was arrived at.  (*d*) Describe measurement, quality and selection of genetic variants.  (*e*) For each exposure, outcome and other relevant variables, describe methods of assessment and diagnostic criteria for diseases.  (*f*) Provide details of ethics committee approval and participant informed consent, if relevant. | Pages 5–8, Supplementary file 1. |
| Assumptions | 5 | Explicitly state the three core IV assumptions for the main analysis (relevance, independence, and exclusion restriction) as well assumptions for any additional or sensitivity analysis. | Page 6, Figure 1 |
| Statistical methods: main analysis | 6 | Describe statistical methods and statistics used.  (*a*) Describe how quantitative variables were handled in the analyses (i.e., scale, units, model).  (*b*) Describe how genetic variants were handled in the analyses and, if applicable, how their weights were selected.  (*c*) Describe the MR estimator (e.g. two-stage least squares, Wald ratio) and related statistics. Detail the included covariates and, in case of two-sample MR, whether the same covariate set was used for adjustment in the two samples.  (*d*) Explain how missing data were addressed.  (*e*) If applicable, say how multiple testing was addressed. | Pages 8–11 |
| Assessment of assumptions | 7 | Describe any methods or prior knowledge used to assess the assumptions or justify their validity. | Pages 8–11 |
| Sensitivity analyses | 8 | Describe any sensitivity analyses or additional analyses performed (e.g. comparison of effect estimates from different approaches, independent replication, bias analytic techniques, validation of instruments, simulations). | Pages 9–11 |
| Software and pre-registration | 9 | (*a*) Name statistical software and package(s), including version and settings used.  (*b*) State whether the study protocol and details were pre-registered (as well as when and where). | Pages 8–11 |
| **Results** |  |  |  |
| Descriptive data | 10 | (*a*) Report the numbers of individuals at each stage of included studies and reasons for exclusion. Consider use of a flow diagram.  (*b*) Report summary statistics for phenotypic exposure(s), outcome(s) and other relevant variables (e.g. means, SDs, proportions).  (*c*) If the data sources include meta-analyses of previous studies, provide the assessments of heterogeneity across these studies.  (*d*) For two-sample MR:  *i.* Provide justification of the similarity of the genetic variant-exposure associations between the exposure and outcome samples.  *ii.* Provide information on the number of individuals who overlap between the exposure and outcome studies. | Not applicable, as we used summary statistics from previously published genome-wide association studies. We cite these accordingly. Furthermore, there is no sample overlap between the exposure and outcome studies. |
| Main results | 11 | (*a*) Report the associations between genetic variant and exposure, and between genetic variant and outcome, preferably on an interpretable scale.  (*b*) Report MR estimates of the relationship between exposure and outcome, and the measures of uncertainty from the MR analysis on an interpretable scale, such as odds ratio or relative risk per SD difference.  (*c*) If relevant, consider translating estimates of relative risk into absolute risk for a meaningful time-period.  (*d*) Consider plots to visualize results (e.g. forest plot, scatterplot of associations between genetic variants and outcome versus between genetic variants and exposure). | Pages 11–16, Table 1, Figure 2, and Supplementary file 2 –  tables and figures |
| Assessment of validity | 12 | (*a*) Report the assessment of the validity of the assumptions.  (*b*) Report any additional statistics (e.g., assessments of heterogeneity across genetic variants, such as I^2^, Q statistic or E-value). | Supplementary file 2 –  tables and figures |
| Sensitivity and additional analyses | 13 | (*a*) Report any sensitivity analyses to assess the robustness of the main results to violations of the assumptions.  (*b*) Report results from other sensitivity analyses or additional analyses.  (*c*) Report any assessment of direction of causal relationship (e.g., bidirectional MR).  (*d*) When relevant, report and compare with estimates from non-MR analyses.  (*e*) Consider any additional plots to visualize results (e.g., leave-one-out analyses). | Pages 16–19 and Supplementary file 2 –  tables and figures. Instrument-risk factor analyses in addition to sensitivity analyses. |
| **Discussion** |  |  |  |
| Key results | 14 | Summarize key results with reference to study objectives. | Page 19 |
| Limitations | 15 | Discuss limitations of the study, taking into account the validity of the IV assumptions, other sources of potential bias, and imprecision. Discuss both direction and magnitude of any potential bias, and any efforts to address them. | Pages 21–22 |
| Interpretation | 16 | (*a*) Meaning: Give a cautious overall interpretation of results in the context of their limitations and in comparison with other studies.  (*b*) Mechanisms: Discuss underlying biological mechanisms that could drive a potential causal relationship between the investigated exposure and the outcome, and whether the gene-environment equivalence assumption is reasonable. Use causal language carefully, clarifying that IV estimates may provide causal effects only under certain assumptions.  (*c*) Clinical relevance: Discuss whether the results have clinical or public policy relevance, and to what extent they inform effect sizes of possible interventions. | Pages 22–23 |
| Generalisability | 17 | Discuss the generalizability of the study results (a) to other populations (i.e. external validity), (b) across other exposure periods/timings, and (c) across other levels of exposure. | Page 21-23 |
| **Other information** |  |  |  |
| Funding | 18 | Describe sources of funding and the role of funders in the present study and, if applicable, sources of funding for the databases and original study or studies on  which the present study is based. | Funding section |
| Data and data sharing | 19 | Provide the data used to perform all analyses or report where and how the data can be accessed, and reference these sources in the article. Provide the statistical code needed to reproduce the results in the article, or report whether the code is publicly accessible and if so, where. | Described, with links provided in the Availability of data and materials section |
| Conflicts of interest | 20 | All authors should declare all potential conflicts of interest. | Page 35 |

Adapted from: BMJ 2021;375:n2233 <http://dx.doi.org/10.1136/bmj.n2233>
